# Application of the WHO Nutrient Profile Model to products on the German market: Implications for proposed new food marketing legislation in Germany

**DOI:** 10.1101/2023.04.24.23288785

**Authors:** Nicole Holliday, Anna Leibinger, Oliver Huizinga, Carmen Klinger, Elochukwu Okanmelu, Karin Geffert, Eva Rehfuess, Peter von Philipsborn

## Abstract

**Background:** Exposure to marketing for foods high in sugar, salt, and fat is considered a key risk factor for childhood obesity. To support efforts to limit such marketing, the World Health Organization Regional Office for Europe has developed a nutrient profile model (WHO NPM). Germany’s Federal Ministry of Food and Agriculture plans to use this model in proposed new legislation on food marketing directed towards children, but the model has not yet been tested on products on the German market. Against this backdrop, the present paper aims to assess the feasibility and implications of implementing the WHO NPM in Germany.

**Methods:** We applied the WHO NPM to a random sample of 660 food and beverage products across 22 product categories on the German market drawn from Open Food Facts, a publicly available product database. We calculated the share of products permitted for marketing to children based on the WHO NPM’s nutrient and ingredient criteria, both under current market conditions and for several hypothetical reformulation scenarios. We also assessed effects of adaptations to and practical challenges in applying the WHO NPM.

**Results:** The median share of products permitted for marketing to children across the model’s 22 product categories was 20% (interquartile range (IQR) 3-59%) and increased to 38% (IQR 11-73%) with model adaptations for fruit juice and milk proposed by Germany’s Federal Ministry of Food and Agriculture. With targeted reformulation (assuming a 30% reduction in fat, sugar, sodium and/or energy) the share of products permitted for marketing to children increased substantially (defined as a relative increase by at least 50%) in several product categories (including bread, processed meat, yogurt and cream, ready-made and convenience foods, and savoury plant-based foods), but changed less in the remaining categories. Practical challenges in applying the model included the ascertainment of the trans-fatty acid content of products, and the classification of products not required to carry nutrient declarations, such as fresh meats, fish, and similar products.

**Discussion:** The application of the WHO NPM to a random sample of food and beverage products on the German market was found to be feasible. Its use in the proposed new legislation on food marketing in Germany seems likely to serve its intended public health objective of limiting marketing in a targeted manner specifically for less healthy products. It seems plausible that it may incentivise reformulation in some product categories. Practical challenges in applying the model could be addressed with appropriate adaptations and procedural provisions.

## Background

There is strong direct and indirect evidence that exposure to marketing for foods high in sugar, salt, and/or fat increases the risk for excess energy intake, unhealthy dietary behaviours, and overweight and obesity among children [1-5]. In many countries, children have been shown to be exposed to extensive marketing for such products [6]. This includes Germany, where children aged 3-13 years consuming media are on average exposed to 15 advertisements for unhealthy foods and beverages per day, including 10 on TV and 5 on the internet [7]. On average, 92% of all food and beverage advertisements to which children are exposed to in Germany are for unhealthy products [7]. This is reflected in the sums spent on food advertisements for different product categories in Germany, which amounted to 1.06 billion € in 2021 for confectionary, compared to 17.2 million € in 2017 for fruit and vegetables (latest available figures, respectively) [8, 9]. Marketing for unhealthy foods and beverages is therefore considered to be an important contributor to the epidemic of obesity and chronic diet-related disease in Germany and internationally [3, 10].

In light of this evidence, Germany’s Federal Ministry of Food and Agriculture published in February 2023 plans for new legislation on food marketing to which children are exposed [11]. The proposed new legislation would limit such marketing to products meeting certain nutrient and ingredient criteria. The law would cover all relevant marketing channels and would use a combination of criteria to define marketing to which children are exposed (see text box 1) [11]. If enacted, the law would be among the most comprehensive regulations of food marketing to which children are exposed worldwide.

### Text box 1

Key parameters of Germany’s proposed new law on food marketing to which children are exposed

Based on the announcements of Germany’s Federal Ministry of Food and Agriculture, the proposed law would be based on the following parameters [11-13]:

- **Nutrient profiling:** The law would limit marketing to products fulfilling the ingredient and nutrient criteria of the second edition of the World Health Organization Regional Office for Europe’s Nutrient Profile Model (WHO NPM) [14], with exceptions for 100% fruit juice (for which marketing would be allowed) and unsweetened milk (for which marketing would be allowed regardless of its fat content) [13].
- **Definition of exposure:** Any advertisement fulfilling at least one of the following three criteria would be considered to be likely to be seen by a substantial number of children, and therefore subject to the law:
  - Any advertisement whose content shows characteristics of being directed towards, or appealing to children (e.g. by using child-like language, comic figures, or children as protagonists).
  - Any advertisement shown in a context in which it is likely to reach an elevated number of children (e.g. during children’s programs on TV, or up to 100 meters around schools, kindergartens, and other facilities frequented by children).
  - Any TV and radio advertisement aired between 6 am and 11 pm.
- **Channels**: The law would cover all relevant marketing channels (including TV, radio, print media, internet including social media and influencer marketing, outdoor advertisement, and sponsoring).
- **Definition of children**: Children are defined as anyone below the age of 14.

The proposed new legislation has been welcomed by medical and health organizations [15, 16], but criticised by food industry groups [17]. Among the more contentious aspects of the proposed legislation is its use of the World Health Organization Regional Office for Europe’s Nutrient Profile Model (WHO NPM) [14]. By some industry groups and other critics of the proposed regulation, the WHO NPM has been criticised for being too far-reaching and impractical, and for amounting to a total ban of advertisement for a broad range of product categories [17-19]. Germany’s Federal Ministry of Food and Agriculture, by contrast, has emphasized that marketing would still be allowed for products qualifying as healthy, and that it expects industry to reformulate products currently not meeting the relevant nutrient and ingredient criteria [11, 12].

As part of its development process, the WHO NPM was pilot-tested in 13 European countries, but not in Germany [14]. Equally, the adaptations to the WHO NPM proposed as part of the planned legislation have not yet been examined systematically. Against this backdrop, the present paper aims to assess the feasibility and implications of implementing the WHO NPM in Germany. Specifically, we apply the WHO NPM (in its original version as well as with the adaptations proposed by Germany’s Federal Ministry of Food and Agriculture) to a random sample of products on the German market in order to assess the following main outcomes:

1. The share of products within each of the WHO NPM’s 22 product categories that meet all its nutrient and ingredient criteria, and which could therefore be marketed to children under the proposed new legislation in Germany.
2. The effects of various hypothetical reformulation scenarios on the share of products meeting the WHO NPM’s nutrient and ingredient criteria.
3. Any practical challenges in applying the WHO NPM to products on the German market.

## Methods

This is a cross-sectional analysis of 660 food and beverage products on the German market randomly sampled from the open source product database Open Food Facts [20]. Our analysis is based on an a priori protocol registered and published with the Open Science Framework [21], and follows the STROBE and STROBE-nut reporting guidelines [22, 23]. In the following sections we provide a short summary of our methodology. Further details are presented in the supplementary material published alongside this manuscript.

### Data sources and methods of assessment

We used product data provided by Open Food Facts, an online open source packaged food and beverage database covering more than 2.8 million products globally, of which slightly more than 200,000 are classified as being available in Germany [20]. We downloaded the complete database, filtered for products classified as being available on the German market, and used R to extract a randomly ordered list of products. We then used the instructions provided by the WHO NPM manual to successively assign these products to the 22 product categories of the WHO NPM until we identified 30 products for each category, or 660 in total [14]. For six product categories that are rare and were therefore not sufficiently represented in the overall sample, we further filtered for relevant keywords (see supplementary material for details). For the 660 included products, we manually extracted nutrient and ingredient information from the Open Food Facts database. The assignment of products to product categories was done by one author (NH, AL, CK, and EO) and double-checked by a second (CK and PvP). Inconsistencies and challenges were discussed in the team of all authors. Data extraction was done by one person (NH, AL, CK, and EO). For a randomly selected sub-sample of all included products, a second author (CK) applied two quality assurance procedures: a) a check if data had been correctly entered from the Open Food Facts database (for 6% of all sampled products), and b) a check if the data provided by the Open Food Facts databased matched nutrient and ingredients data provided on the websites of manufacturers or (if no manufacturer website could be identified) on the websites of online retailers (for 10% of all sampled products). Overall, we found the data provided by Open Food Facts to be reasonably reliable (see supplementary material for further details).

### Main variables

We calculated the following variables for each of the 22 product categories defined by the WHO NPM:

- The share of products not exceeding any nutrient or ingredient threshold (i.e. products permitted for marketing to children under the WHO NPM).
- The share of products not exceeding specific nutrient or ingredient thresholds (of those products to which the respective threshold applies) (e.g. the share of products not exceeding the total fat threshold, of all products for which the WHO NPM defines a total fat threshold).
- The share of products for each of the 22 product categories that exceed 0, 1, 2, or 3 thresholds and the mean number of thresholds exceeded in each category by those products that exceed at least one threshold.

The analysis was conducted separately for the original WHO NPM and for the WHO NPM with the adaptations proposed by Germany’s Federal Ministry of Food and Agriculture. We used the nutrient and ingredient thresholds as described in the WHO NPM manual (see table 1), except for the trans-fatty acid threshold, which we were unable to assess due to a lack of publicly available data on the trans-fatty acid content of products on the German market (for further details see the section on challenges below) [14]. All analyses were performed in R (the R code is shown in the supplementary material). We used the conditional formatting function of MS Excel to create color-coded tables for illustration purposes. In this color-coding, shades of green stand for values that are desirable from a public health-perspective (e.g. a high share of products not exceeding nutrient or ingredient thresholds); shades of red stand for less desirable values, and shades of yellow for values that are in-between.

**Table 1:** Nutrient and ingredient thresholds of the WHO NPM [14].

| **Table 1: Nutrient and ingredient thresholds of the WHO NPM [14]** |  |  |  |  |  |  |  |  |
| --- | --- | --- | --- | --- | --- | --- | --- | --- |
| **Nr** | **Product category\*** | **Energy (kcal)** | **Total fat (g)** | **Saturated fat (g)** | **Total sugars (g)** | **Sodium (g)** | **Added sugars (g)** | **Non-sugar sweeteners (g)** |
| **1** | Confectionery |  |  |  |  |  | 0 | 0 |
| **2** | Cakes and cookies |  | 3 |  |  | 0.1 | 0 | 0 |
| **3** | Savoury snacks |  |  |  |  | 0.1 | 0 | 0 |
| **4** | Beverages |  |  |  |  |  |  |  |
| **4.1** | Juices |  |  |  | 0** |  |  | 0 |
| **4.2** | Dairy milk drinks |  | 3** |  |  |  | 0 | 0 |
| **4.3** | Plant-based milks |  | 3 |  |  |  | 0 | 0 |
| **4.4** | Energy drinks |  |  |  |  |  | 0 | 0 |
| **4.5** | Soft drinks, bottled water and other drinks |  |  |  |  |  | 0 | 0 |
| **5** | Ice cream |  | 3 |  |  | 0.1 | 0 | 0 |
| **6** | Breakfast cereals |  | 17 |  | 12.5 | 0.5 |  |  |
| **7** | Yogurt and cream |  | 3 | 1 | 12.5 | 0.1 |  |  |
| **8** | Cheese |  | 17 |  |  | 0.5 |  |  |
| **9** | Ready-made and convenience foods | 225 | 17 | 6 | 12.5 | 0.5 |  |  |
| **10** | Butter, other fats and oils |  |  | 21 |  | 0.5 |  |  |
| **11** | Bread |  | 17 |  | 12.5 | 0.5 |  |  |
| **12** | Pasta and grains |  | 17 |  | 12.5 | 0.5 |  |  |
| **13** | Fresh and frozen meat, fish and eggs |  | 17 |  |  |  |  |  |
| **14** | Processed meat and fish |  | 17 |  |  | 0.5 |  |  |
| **15** | Fresh and frozen fruit and vegetables | Permitted |  |  |  |  |  |  |
| **16** | Processed fruit and vegetables |  | 3 |  | 12.5 | 0.5 | 0 |  |
| **17** | Savoury plant-based foods |  | 17 |  |  | 0.5 | 0 | 0 |
| **18** | Sauces, dips and dressings |  | 17 |  |  | 0.5 | 0 | 0 |
| Abbreviations: WHO NPM: World Health Organization Regional Office for Europe Nutrient Profiling Model [14]. *A detailed description of each category, including examples, is provided in the supplementary material. ** These thresholds apply only in the original WHO NPM, and not in the adapted version proposed by Germany's Federal Ministry of Food Agriculture in the context of the model's planned used in Germany. Source: [13, 14] |  |  |  |  |  |  |  |  |

### Hypothetical reformulation scenarios

We also assessed how several hypothetical reformulation scenarios would affect the variables listed above. We defined these reformulation scenarios based on two considerations, as described in our study protocol [21]: i) the nutrient or ingredient thresholds most commonly exceeded in the respective product category, and ii) existing reformulation commitments made by industry groups as part of Germany’s National Strategy for the Reduction of Sugar, Salt and Fat in Processed Foods [24].

### Feasibility of applying the WHO NPM

During the research process, all authors individually took note of any difficulties or challenges in any of the steps involved in applying the WHO NPM. These were subsequently deliberated and structured by the team in several rounds of discussions. Based on these, one author (NH) drafted a list of challenges, which were again discussed by all authors, and subsequently revised by a second author (PvP) and double-checked by all remaining authors.

## Results

### Share of products permitted for marketing to children

The median share of products across the 22 product categories that meet all the nutrient and ingredient criteria and are therefore permited for marketing to children is 20% (interquarle range (IQR) 3%-59%) under the WHO NPM (see table 2) [14]. With the adaptaons to the WHO NPM proposed by Germany’s Federal Ministry of Food and Agriculture in the context of the model’s planned use in Germany, this share increases to 38% (IQR 11%-73%). These adaptaons increase the share of products permited for marketing to children from 0% to 100% in the category of juices (by removing the threshold for total sugars), and from 20% to 80% in the category of milk (by removing the threshold for total fat).

**Table 2:**
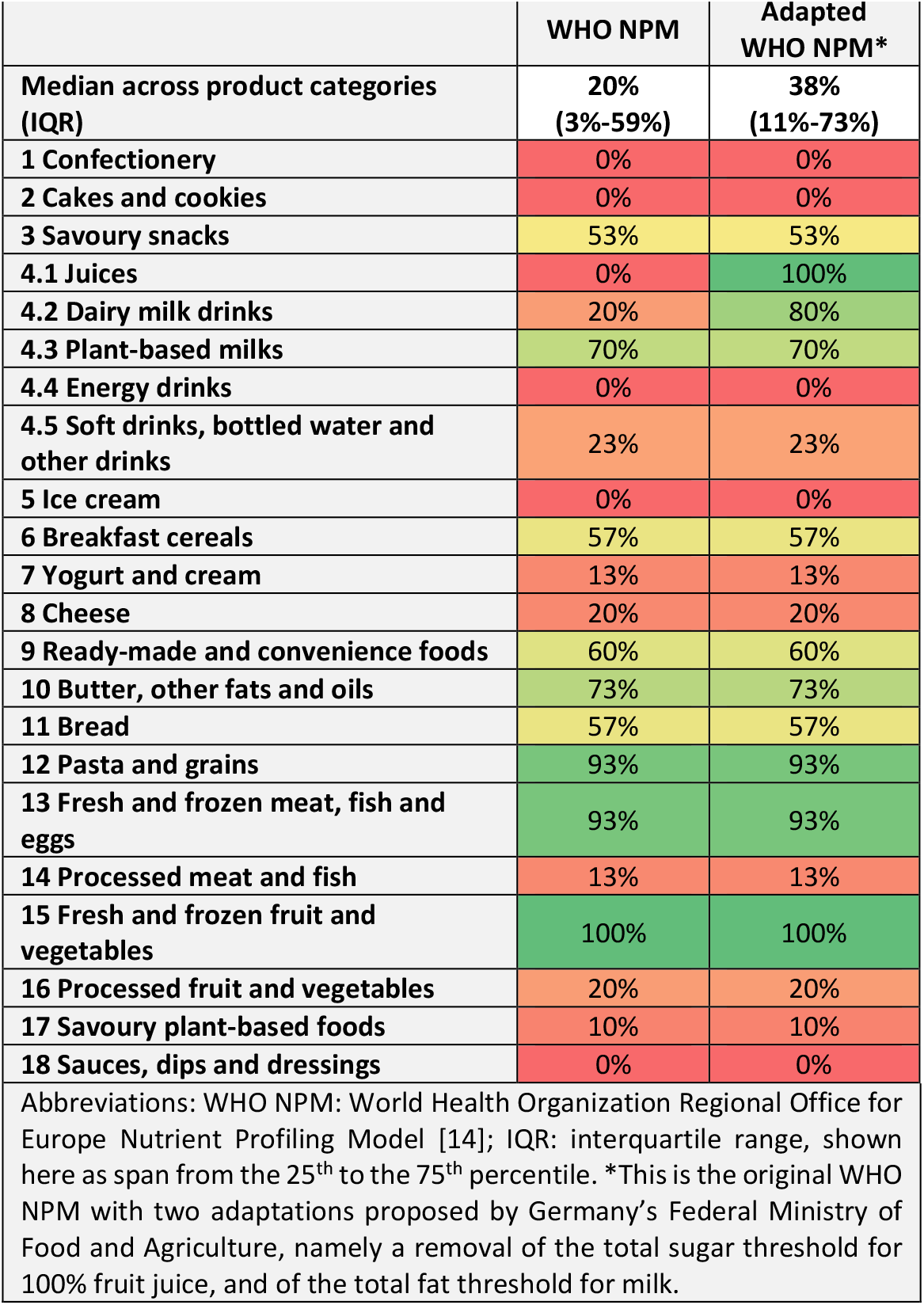
Share of products meeting all nutrient and ingredient criteria of the WHO NPM (i.e. permitted for marketing to children)

|  |  |  |
| --- | --- | --- |
|  | WHO NPM | Adapted WHO NPM* |
| Median across product categories (IQR) | 20%<br>(3%-59%) | 38%<br>(11%-73%) |
| 1 Confectionery | 0% | 0% |
| 2 Cakes and cookies | 0% | 0% |
| 3 Savoury snacks | 53% | 53% |
| 4.1 Juices | 0% | 100% |
| 4.2 Dairy milk drinks | 20% | 80% |
| 4.3 Plant-based milks | 70% | 70% |
| 4.4 Energy drinks | 0% | 0% |
| 4.5 Soft drinks, bottled water and other drinks | 23% | 23% |
| 5 Ice cream | 0% | 0% |
| 6 Breakfast cereals | 57% | 57% |
| 7 Yogurt and cream | 13% | 13% |
| 8 Cheese | 20% | 20% |
| 9 Ready-made and convenience foods | 60% | 60% |
| 10 Butter, other fats and oils | 73% | 73% |
| 11 Bread | 57% | 57% |
| 12 Pasta and grains | 93% | 93% |
| 13 Fresh and frozen meat, fish and eggs | 93% | 93% |
| 14 Processed meat and fish | 13% | 13% |
| 15 Fresh and frozen fruit and vegetables | 100% | 100% |
| 16 Processed fruit and vegetables | 20% | 20% |
| 17 Savoury plant-based foods | 10% | 10% |
| 18 Sauces, dips and dressings | 0% | 0% |
| Abbreviations: WHO NPM: World Health Organization Regional Office for Europe Nutrient Profiling Model [14]; IQR: interquartile range, shown here as span from the 25<sup>th</sup> to the 75<sup>th</sup> percentile. *This is the original WHO NPM with two adaptations proposed by Germany's Federal Ministry of Food and Agriculture, namely a removal of the total sugar threshold for 100% fruit juice, and of the total fat threshold for milk. |  |  |

### Share of products meeting specific nutrient and ingredient criteria

Table 3 shows the share of products meeng the respective criterion for each of the nutrient and ingredient criteria of the adapted WHO NPM. This share ranges from 100% (e.g., all breakfast cereals in our sample are below the total fat threshold for that product category) to 0% (e.g., none of the cakes and cookies in our sample meets the WHO NPM’s criterion of not containing added sugars). Across the 22 product categories, the criterion that was most commonly met was that products may not contain non-sugar sweeteners (median 100%, IQR 97%-100%). In most product categories for which this criterion applies, all or almost all of the products in our sample did not contain any non-sugar sweeteners, and therefore met this criterion (the only excepons are energy drinks, as well as so drinks, botled water and other drinks). By contrast, the criterion most rarely met was the one that products may not contain added sugars (median 37%, IQR 10%-58%): in most product categories to which this criterion applies, a majority of products did not meet it (see table 3).

**Table 3:**
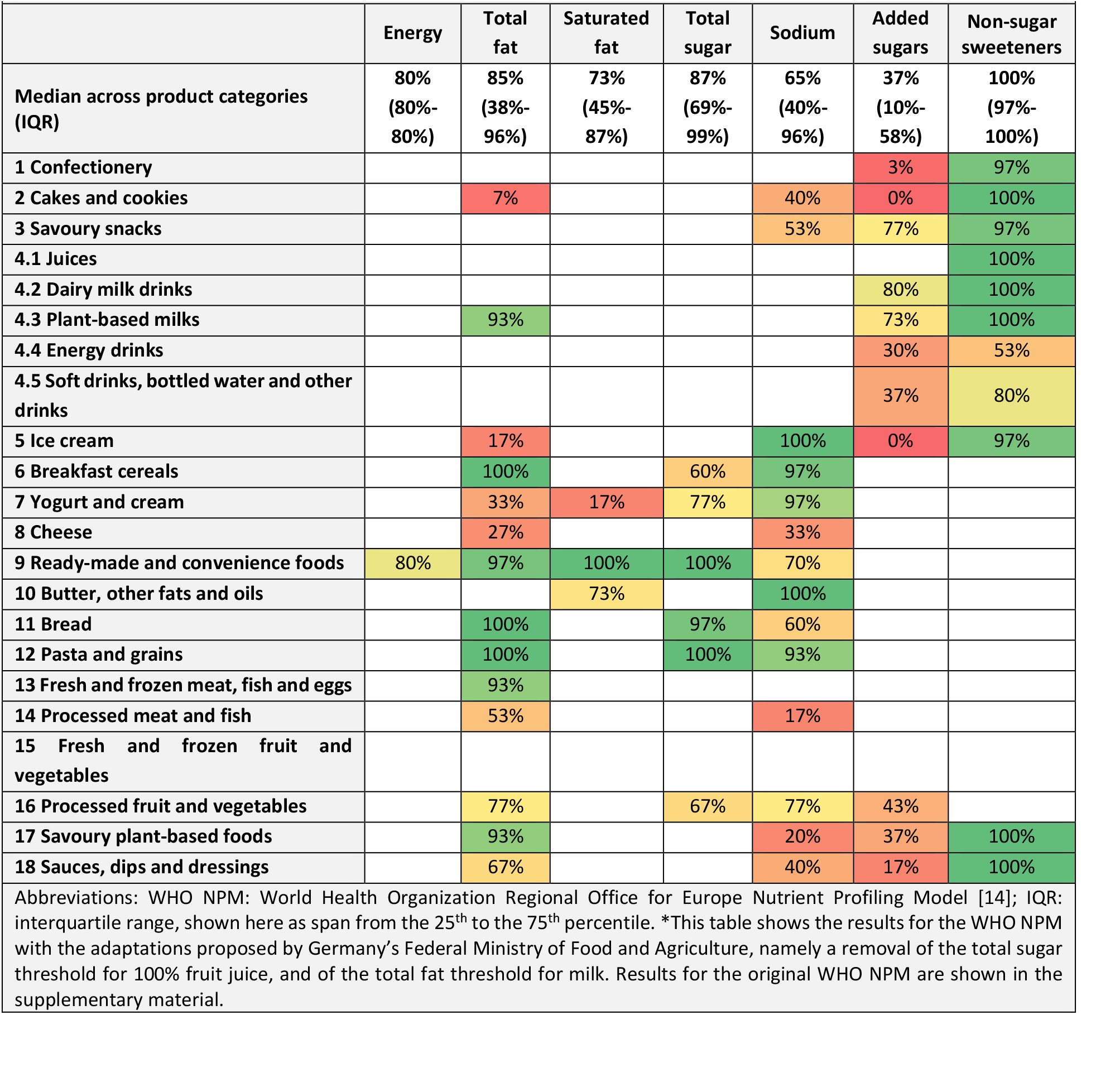
Share of products meeting the adapted WHO NPM’s nutrient and ingredient criteria*.

|  |  |  |  |  |  |  |  |
| --- | --- | --- | --- | --- | --- | --- | --- |
|  | Energy | Total fat | Saturated fat | Total sugar | Sodium | Added sugars | Non-sugar sweeteners |
| Median across product categories (IQR) | 80%<br>(80%-80%) | 85%<br>(38%-96%) | 73%<br>(45%-87%) | 87%<br>(69%-99%) | 65%<br>(40%-96%) | 37%<br>(10%-58%) | 100%<br>(97%-100%) |
| 1 Confectionery |  |  |  |  |  | 3% | 97% |
| 2 Cakes and cookies |  | 7% |  |  | 40% | 0% | 100% |
| 3 Savoury snacks |  |  |  |  | 53% | 77% | 97% |
| 4.1 Juices |  |  |  |  |  |  | 100% |
| 4.2 Dairy milk drinks |  |  |  |  |  | 80% | 100% |
| 4.3 Plant-based milks |  | 93% |  |  |  | 73% | 100% |
| 4.4 Energy drinks |  |  |  |  |  | 30% | 53% |
| 4.5 Soft drinks, bottled water and other drinks |  |  |  |  |  | 37% | 80% |
| 5 Ice cream |  | 17% |  |  | 100% | 0% | 97% |
| 6 Breakfast cereals |  | 100% |  | 60% | 97% |  |  |
| 7 Yogurt and cream |  | 33% | 17% | 77% | 97% |  |  |
| 8 Cheese |  | 27% |  |  | 33% |  |  |
| 9 Ready-made and convenience foods | 80% | 97% | 100% | 100% | 70% |  |  |
| 10 Butter, other fats and oils |  |  | 73% |  | 100% |  |  |
| 11 Bread |  | 100% |  | 97% | 60% |  |  |
| 12 Pasta and grains |  | 100% |  | 100% | 93% |  |  |
| 13 Fresh and frozen meat, fish and eggs |  | 93% |  |  |  |  |  |
| 14 Processed meat and fish |  | 53% |  |  | 17% |  |  |
| 15 Fresh and frozen fruit and vegetables |  |  |  |  |  |  |  |
| 16 Processed fruit and vegetables |  | 77% |  | 67% | 77% | 43% |  |
| 17 Savoury plant-based foods |  | 93% |  |  | 20% | 37% | 100% |
| 18 Sauces, dips and dressings |  | 67% |  |  | 40% | 17% | 100% |

Abbreviations: WHO NPM: World Health Organization Regional Office for Europe Nutrient Profiling Model [14]; IQR: interquartile range, shown here as span from the 25<sup>th</sup> to the 75<sup>th</sup> percentile. \*This table shows the results for the WHO NPM with the adaptations proposed by Germany's Federal Ministry of Food and Agriculture, namely a removal of the total sugar threshold for 100% fruit juice, and of the total fat threshold for milk. Results for the original WHO NPM are shown in the supplementary material.

### Average nutrient content

Table 4 shows the median nutrient content of products relative to the respective threshold. In most product categories, the median content of energy, fat, sugar, and salt is below the relevant threshold defined by the WHO NPM. For example, the median content of total sugars in breakfast cereals in our sample stands at 84% of the total sugars threshold of the WHO NPM (which is 12.5 g/100g, see table 2). By contrast, the total fat content of cakes and cookies stands at 700% of the WHO NPM’s threshold (which is 3 g/100 g).

**Table 4:**
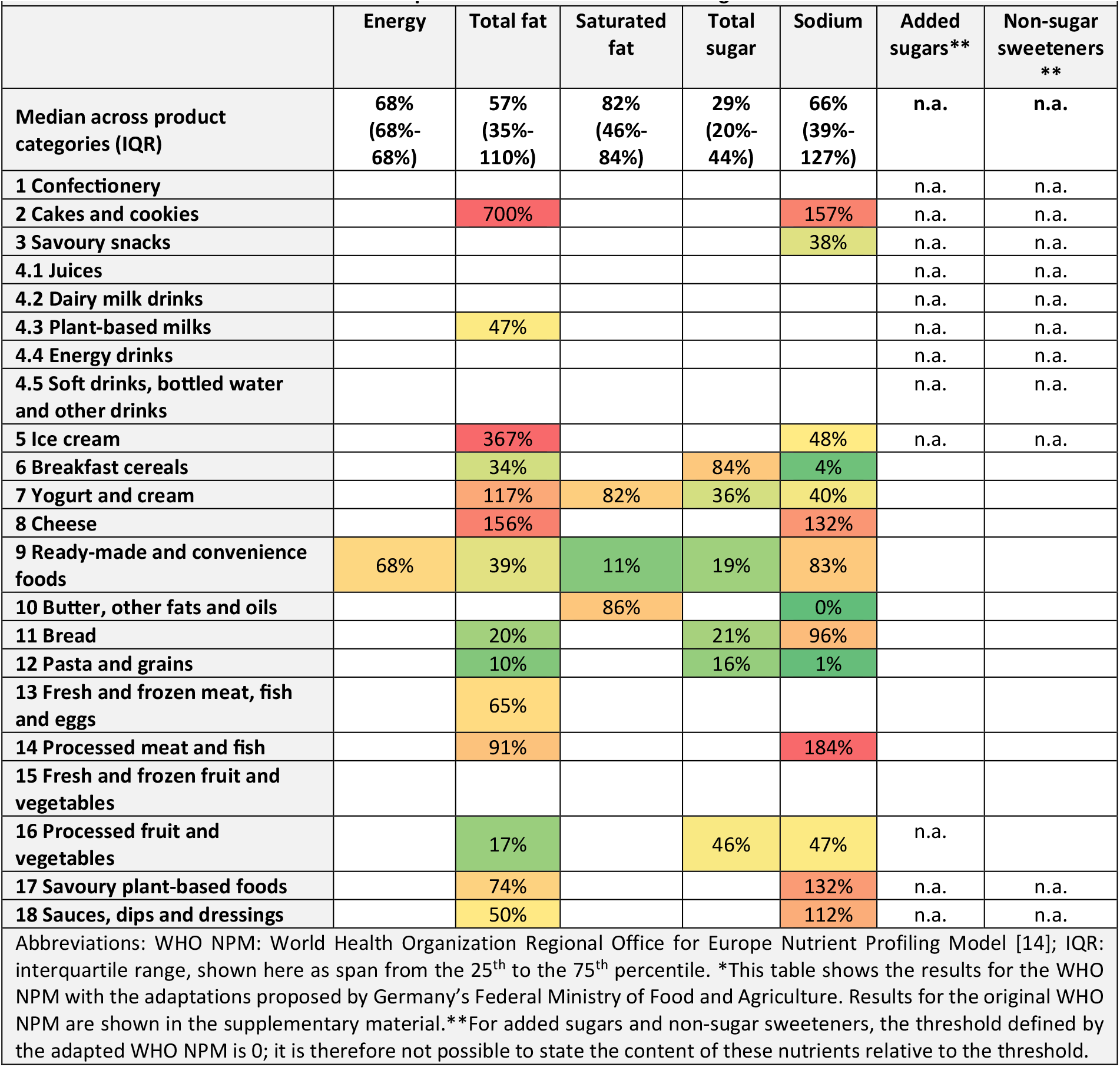
Median content relative to the adapted WHO NPM’s nutrient and ingredient thresholds*.

### Number of thresholds exceeded

In most of the product categories, a majority of products either do not exceed any threshold, or exceed only one threshold (median shares of 38% and 27%, respectively, see table 5). Consequently, in most of the product categories, the share of products exceeding two thresholds (median 7%) or even three thresholds (median 0%) is low. No product in our sample exceeded more than three thresholds. In only one product category was there a majority of products that exceeded three thresholds: among the cakes and cookies in our sample, 60% of products exceeded three thresholds (mostly those for total fat, sodium, and added sugars).

**Table 5:**
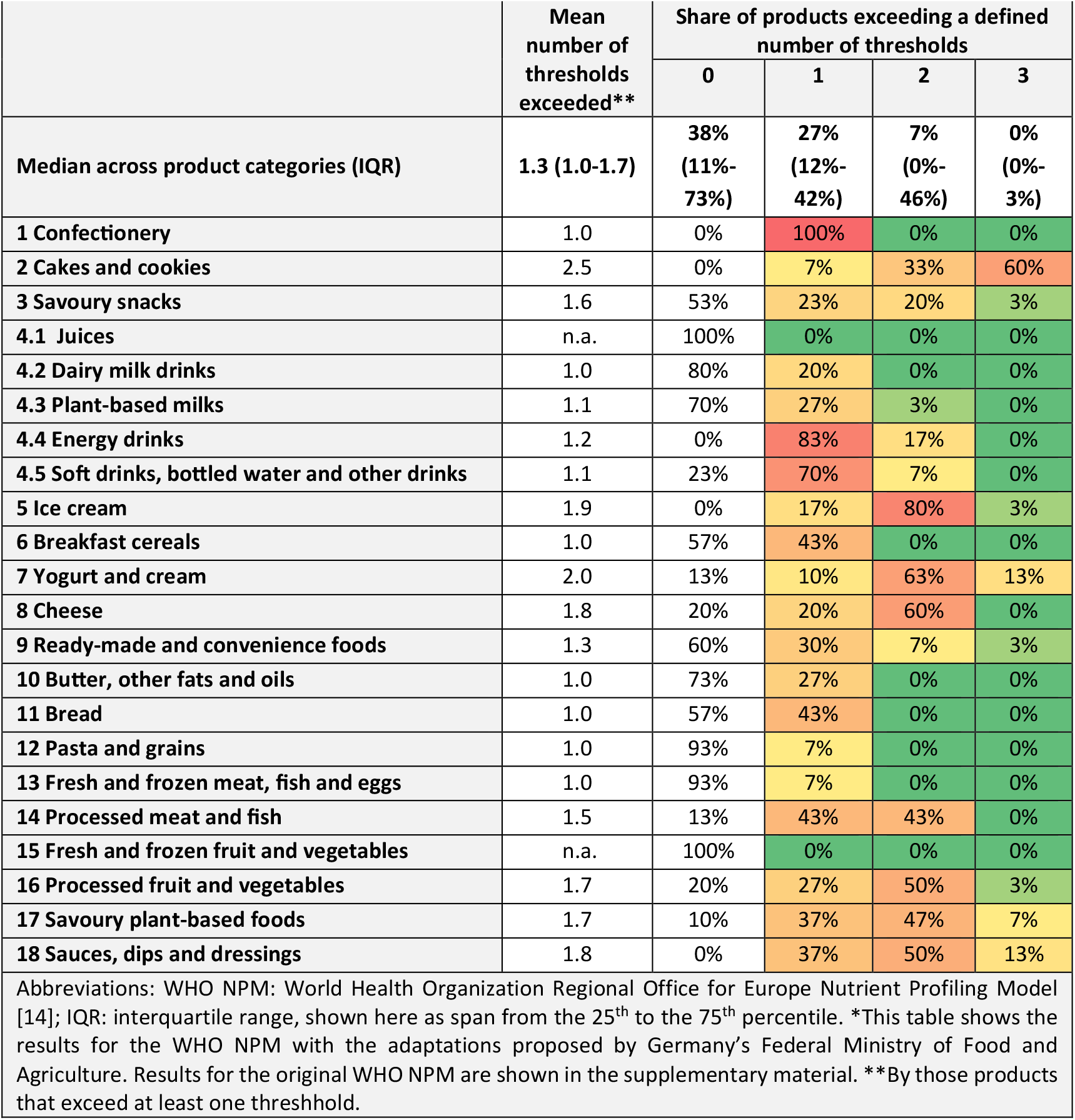
Share of products exceeding a defined number of thresholds (WHO NPM with adaptation)*

|  | Mean number of thresholds exceeded** | Share of products exceeding a defined number of thresholds |  |  |  |
| --- | --- | --- | --- | --- | --- |
|  |  | 0 | 1 | 2 | 3 |
| **Median across product categories (IQR)** | **1.3 (1.0-1.7)** | **38%**<br>**(11%-73%)** | **27%**<br>**(12%-42%)** | **7%**<br>**(0%-46%)** | **0%**<br>**(0%-3%)** |
| **1 Confectionery** | 1.0 | 0% | 100% | 0% | 0% |
| **2 Cakes and cookies** | 2.5 | 0% | 7% | 33% | 60% |
| **3 Savoury snacks** | 1.6 | 53% | 23% | 20% | 3% |
| **4.1 Juices** | n.a. | 100% | 0% | 0% | 0% |
| **4.2 Dairy milk drinks** | 1.0 | 80% | 20% | 0% | 0% |
| **4.3 Plant-based milks** | 1.1 | 70% | 27% | 3% | 0% |
| **4.4 Energy drinks** | 1.2 | 0% | 83% | 17% | 0% |
| **4.5 Soft drinks, bottled water and other drinks** | 1.1 | 23% | 70% | 7% | 0% |
| **5 Ice cream** | 1.9 | 0% | 17% | 80% | 3% |
| **6 Breakfast cereals** | 1.0 | 57% | 43% | 0% | 0% |
| **7 Yogurt and cream** | 2.0 | 13% | 10% | 63% | 13% |
| **8 Cheese** | 1.8 | 20% | 20% | 60% | 0% |
| **9 Ready-made and convenience foods** | 1.3 | 60% | 30% | 7% | 3% |
| **10 Butter, other fats and oils** | 1.0 | 73% | 27% | 0% | 0% |
| **11 Bread** | 1.0 | 57% | 43% | 0% | 0% |
| **12 Pasta and grains** | 1.0 | 93% | 7% | 0% | 0% |
| **13 Fresh and frozen meat, fish and eggs** | 1.0 | 93% | 7% | 0% | 0% |
| **14 Processed meat and fish** | 1.5 | 13% | 43% | 43% | 0% |
| **15 Fresh and frozen fruit and vegetables** | n.a. | 100% | 0% | 0% | 0% |
| **16 Processed fruit and vegetables** | 1.7 | 20% | 27% | 50% | 3% |
| **17 Savoury plant-based foods** | 1.7 | 10% | 37% | 47% | 7% |
| **18 Sauces, dips and dressings** | 1.8 | 0% | 37% | 50% | 13% |
| Abbreviations: WHO NPM: World Health Organization Regional Office for Europe Nutrient Profiling Model [14]; IQR: interquartile range, shown here as span from the 25<sup>th</sup> to the 75<sup>th</sup> percentile. *This table shows the results for the WHO NPM with the adaptations proposed by Germany's Federal Ministry of Food and Agriculture. Results for the original WHO NPM are shown in the supplementary material. **By those products that exceed at least one threshold. |  |  |  |  |  |

### Specific reformulation scenarios

We defined 15 reformulation scenarios for 8 product categories (see table 6). In some product categories, a moderate reduction in a single nutrient is sufficient to substantially increase the share of products permitted for marketing to children under the WHO NPM. For example, a reduction of the sodium content of bread by 20% raises the share of products permitted for marketing to children in this category from 57% to 90%. In other product categories, the effects of reformulation are less pronounced; e.g., for cereals, a reduction in the content of total sugars by 30% increases the share of products permitted for marketing to children from 57% to 67% (see table 6).

**Table 6:** Effects of specific reformulation scenarios on the share of products permited for marketing to children (in %)

| Table 6: Effects of specific reformulation scenarios on the share of products permitted for marketing to children (in %) |  |  |  |  |  |  |
| --- | --- | --- | --- | --- | --- | --- |
| Category | Threshold | Reformulation level |  |  |  | Relative increase* |
|  |  | 0% | 10% | 20% | 30% |  |
| Bread | Sodium | 57% | 73% | 90% | 90% | 58% |
| Breakfast cereals | Sugar | 57% | 57% | 63% | 67% | 17% |
| Ready-made and convenience foods | Sodium | 60% | 63% | 73% | 80% | 33% |
|  | Sodium and energy | 60% | 70% | 83% | 90% | 50% |
| Savoury plant-based food | Sodium | 10% | 20% | 23% | 27% | 167% |
| Processed meat and fish | Sodium | 13% | 17% | 17% | 17% | 28% |
|  | Total fat and sodium | 13% | 20% | 23% | 23% | 77% |
| Savoury snacks | Sodium | 53% | 53% | 57% | 57% | 7% |
| Yogurt and cream | Saturated fat | 13% | 17% | 20% | 20% | 50% |
|  | Saturated fat and sugar | 13% | 20% | 23% | 23% | 75% |
|  | Saturated fat and total fat | 13% | 17% | 20% | 20% | 50% |
|  | Saturated fat, total fat, and total sugar | 13% | 20% | 23% | 23% | 75% |
| Cheese | Sodium | 20% | 20% | 20% | 20% | 0% |
|  | Total fat | 20% | 20% | 20% | 20% | 0% |
|  | Total fat and sodium | 20% | 20% | 20% | 27% | 35% |
| *Relative increase between the baseline (no reformulation) and the -30% reformulation scenario. Note: This table shows the results for the WHO NPM with the adaptations proposed by Germany's Federal Ministry of Food and Agriculture, namely a removal of the total sugar threshold for 100% fruit juice, and of the total fat threshold for milk. Results for the original WHO NPM are shown in the supplementary material. |  |  |  |  |  |  |

### Experiences in applying the WHO NPM and challenges encountered

We encountered three main challenges in applying the WHO NPM to our sample of products from the German market. Specifically, we found that for some products and some nutrient and ingredient criteria, the nutrient and ingredient information provided on the packages of food and beverage products in Germany is not sufficient to determine if the respective criterion is met or not. Specifically, this refers to: i) the trans-fatty acid content of products; ii) the nutrient content of products currently not required to display nutrient declarations on their packages (such as fresh meat); and iii) the nutrient content of products prepared by the consumer with dry mixes, powders, or concentrates. A detailed description of these challenges and how we addressed them in our analysis is provided in table 7. We also encountered a number of challenges related to the assignment of products to product categories, which are described in the supplementary material.

**Table 7:**
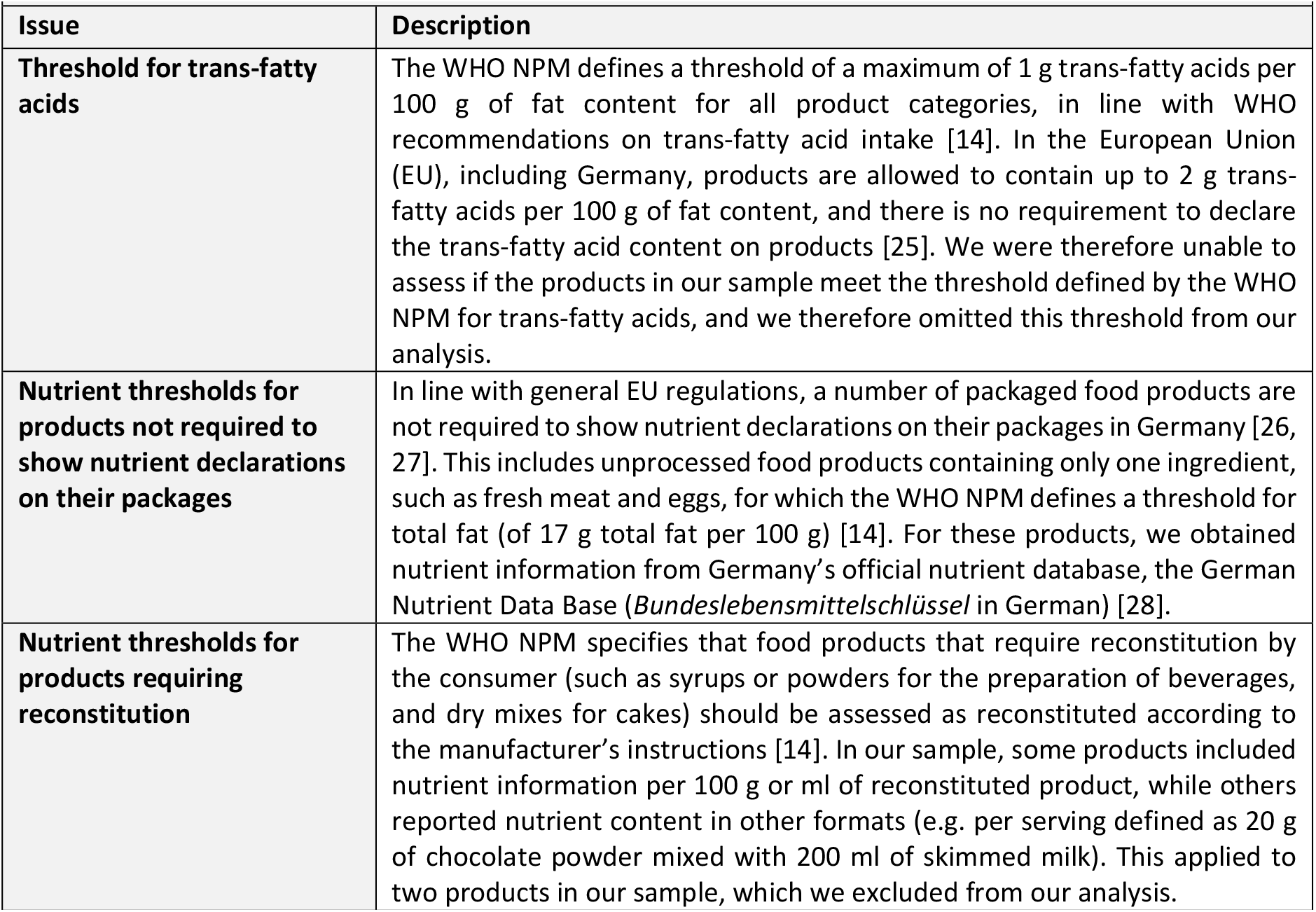
Challenges in applying the WHO NPM to products on the German market.

## Discussion

### Key findings

Applying the WHO NPM as proposed by the WHO Regional Office for Europe to our random sample of 660 food and beverage products on the German market, the median share of products across the model’s 22 product categories that meet all its nutrient and ingredient criteria, and would therefore be permitted for marketing to children is 20% (interquartile range (IQR) 3%-59%). With the introduction of model adaptations announced by Germany’s Federal Ministry of Food and Agriculture in the context of the model’s planned use in Germany, this figure increases to 38% (IQR 11%-73%). The product categories in which the highest share of products meet all criteria of the adapted WHO NPM are fresh and frozen fruit and vegetables (100% of all products in our sample), fruit juices (100%), fresh and frozen meat, fish and eggs (93%), pasta and grains (93%) and dairy milk drinks (80%). By contrast, none of the products in our sample met the WHO NPM’s nutrient and ingredient criteria in the categories of confectionery, cakes and cookies, energy drinks, ice cream, and sauces, dips and dressings.

Most products that exceed at least one threshold defined by the WHO NPM (and are therefore not permited for marketing to children) exceed only one of the model’s seven nutrient and ingredient thresholds (the median across the 22 product categories of the mean number of thresholds exceeded is 1.3). This suggests that in many cases, a targeted reduction in only one or two nutrients of concern would be sufficient to raise the share of products permited for marketing to children. Indeed, with targeted reformulation (assuming a 30% reduction in fat, sugar, sodium, and/or energy) the share of products permitted for marketing to children increases substantially (defined as a relative increase by at least 50%) in several product categories (including bread, processed meat, yogurt and cream, ready-made and convenience foods, and savoury plant-based foods). In other product categories, however, the effects of reformulation are less pronounced. This applies in particular to product categories for which the WHO NPM defines a threshold for added sugars of zero (this includes confectionery, cakes and cookies, energy drinks, soft drinks, botled water and other drinks,, ice cream, processed fruit and vegetables, and sauces, dips and dressings); in these product categories, it would be necessary to replace added sugars completely with other ingredients to increase the share of products permited for marketing to children substantially. Indeed, the added sugars threshold is the WHO NPM’s most commonly exceeded threshold, and the only threshold for which more than half of all products in most product categories are above the threshold defined by the WHO NPM. By contrast, for all other nutrient and ingredient thresholds, in most product categories a majority of products are below the respective threshold.

### Strengths and limitations

Our study has several strengths. It is, to the best of our knowledge, the first study applying the WHO NPM to a sample of products from the German market. We examine a number of questions of critical relevance to the ongoing policy-making process regarding the proposed new legislation regulating food and beverage marketing directed towards children in Germany [11]. Our study is based on a random sample of products from Open Food Facts, a large, publicly available food and beverage database, and we implemented a number of quality assurance procedures. Our study follows the STROBE and STROBE-nut reporting guidelines [22, 23], and is based on an a priori protocol, which we developed and registered publicly before we conducted our analyses [21]. Finally, our manuscript’s supplementary material includes a detailed description of methods, data, and code, ensuring transparency and allowing for the reproducibility of our research.

Besides these strengths, our study also has a number of limitations. The products contained in the Open Food Facts database are not necessarily representative of the German market overall, and we were unable to account for the market share of different products (e.g. by conducting separate analyses for top-selling products). While we found the nutrient and ingredient data provided by Open Food Facts to be reasonably reliable, our spot checks revealed that for some products, data differed from the product data provided on the manufacturers’ and retailers’ websites, indicating that for some products Open Food Facts may provide outdated information. Moreover, due to resource limitations, our analysis was limited to 30 products per category, and a larger number of products may have yielded results with a higher precision.

### Policy implications

Overall, we found the application of the WHO NPM to products on the German market to be feasible. We encountered only minor practical challenges, which could be addressed by appropriate adaptations and procedural provisions. Specifically, an application of the threshold for trans-fatty acids would be challenging in Germany unless new rules on the declaration of trans-fatty acids were introduced; this would, however, require changes to EU regulation [27]. Moreover, provisions are needed to determine the nutrient content of products that are currently not required to show nutrient declarations on their packages (such as fresh, unprocessed meat, fish, and similar products) [26, 27]. The same applies for the nutrient content of foods prepared by the final consumer from concentrates or dry mixes (such as beverage powders and baking mixes). In some cases, more detailed guidance on the assignment of products to product categories would be helpful.

Regarding milk, Germany’s Federal Ministry of Food and Agriculture announced that it is planning to adapt the WHO NPM by not applying its threshold for total fat for this product category (which is 3 g/100 ml), thus allowing marketing for milk with a higher fat content [13]. In our analysis, this adaptation increased the share of permitted products in the respective category of the WHO NPM (dairy milk drinks) considerably, from 20% to 80%. For consistency reasons, it may be worth considering extending this adaptation to category 4.3 (plant-based milks), and to category 7 (yogurt, sour milk, cream and similar products), for which the WHO NPM currently also defines thresholds for total fat of 3 g per 100 ml (for yogurt and similar products it additionally defines a threshold for saturated fat of 1 g per 100 ml) [14]. In our sample of products, a removal of these thresholds would raise the share of products permitted for marketing to children in the category of plant-based milks from 70% to 73%, and in the category of yogurt, sour milk, cream and similar products from 13% to 73%.

Regarding the stringency of the WHO NPM, we found that in most product categories, a substantial number of products in our sample met all nutrient and ingredients criteria defined by the WHO NPM and could still be advertised to children under Germany’s proposed new food marketing regulation. This suggests that the proposed new legislation would not result in a total ban of food marketing (as claimed by some critics [17, 19]), and that food marketing efforts could be redirected towards existing products with a lower content of sugar, salt, and fat, as intended by Germany’ Federal Ministry of Food and Agriculture [12].

Our analysis of reformulation scenarios shows that in several product categories, the share of permitted products could be increased substantially through moderate reductions in the content of fat, sugar, salt, and/or energy. This suggests that the use of the WHO NPM in the proposed new marketing regulation in Germany could indeed provide incentives for reformulation, as hoped for by Germany’s Federal Ministry of Food and Agriculture [12].

## Conclusions

The application of the WHO NPM to food and beverage products on the German market was found to be feasible. Challenges encountered in the application of the WHO NPM in its current form in Germany could be addressed with appropriate adaptations and provisions. The share of products that meet all criteria defined by the WHO NPM and are therefore permitted for marketing to children under the model varies considerably by product category. In most, but not all product categories, this share is substantial. In several product categories, the share of products permitted for marketing to children could be increased substantially with targeted reformulation. The use of the WHO NPM in the context of the proposed new legislation on food marketing in Germany seems feasible. It seems plausible that it will serve its intended public health objective of limiting marketing in a targeted manner specifically for less healthy products, and possibly also of incentivising reformulation in some product categories.

## Data Availability

The data used in this research is available from the authors upon reasonable request.

## Statements

## Acknowledgements

We thank Cody Holliday for their generous support with data management and analysis.

## Statement of Ethics

No human subjects were involved in this research and no ethical clearance was sought in line with the regulations of the Ethics Committee of Ludwig-Maximilians-Universität München (LMU Munich).

## Author Contributions

Conceptualization: PvP and OH; Methodology: PvP, OH, NH, AL, CK, ER; Investigation: NH, AL, PvP, CK, EO, KG; Data Curation: NH and AL; Formal Analysis: AL and NH; Writing – Original Draft: PvP and NH; Writing – Review and Editing: NH, AL, OH, CK, EO, KG, ER, PvP; Supervision: PvP.

## Study registration and protocol availability and fidelity

Registered with Open Science Framework (OSF) after data was collected, but before analyses were conducted at https://osf.io/bjevc [21]. Any discrepancies between the protocol and the study have been explained in the the supplementary material.

## Data Availability Statement

The data used in this research is available from the authors upon reasonable request.

## Supplementary material

Additional information is provided in the supplementary material published online alongside this article.

