## Supplementary material for "Application of the WHO Nutrient Profile Model to products on the German market: Implications for proposed new food marketing legislation in Germany"

**Table of content**

1. **Detailed description of the methodology** ... 1
 1.1. **Sampling and data extraction** ... 1
 1.2. **Quality control procedures** ... 3
 1.3. **Assignment of products to product categories** ... 4
2. **Detailed description of the product categories defined by the WHO NPM** ... 6
3. **Additional analyses** ... 9
4. **Differences between protocol and manuscript** ... 11
5. **Code** ... 12
 5.1. **Code for the random sampling (for R)** ... 12
 5.2. **Code for the statistical analyses (for R)** ... 13
6. **Sampling and data extraction guidance sheet** ... 24
7. **STROBE-nut reporting guideline** ... 27
8. **List of products excluded after reassignment** ... 34
**References** ... 38

### 1. Detailed description of the methodology

#### 1.1. Sampling and data extraction

***Sampling procedure***

The data on food and beverage products on the German market was sourced from Open Food Facts [1]. Open Food Facts is, to the best of the authors' knowledge, the largest, open-source, and publicly available database of food and beverage products on the German market [1]. We downloaded the entire data set from the global version of Open Food Facts and then used the software RipGrep to filter for all entries containing the string "en: Germany" to filter for products that were tagged in Open Food Facts as being sold in Germany [2]. The resulting smaller data set was used for analysis and saved as a JSON file. We then used the software JQ to extract the product name and brand (where applicable) from the JSON file and transform it into a CSV file [3]. We then imported this CSV file into R, where we randomly sampled products in batches of 50 or 100 at a time (see section 4.1 for the R script used for this purpose) [4]. One author (NH, AL, CK, or EO) then successively checked if these products fell within the scope of the WHO NPM, and if the Open Food Facts entry for the respective product was complete (i.e. provided all the information needed for our data extraction, see below), and assigned all eligible products to the corresponding product category of the WHO NPM, following the instructions provided in the WHO NPM manual [5]. To support consistency among reviewers, we created a sampling and data extraction guidance sheet (see section 5).

Products were randomly sampled according to this method until 16 of the 22 categories had been filled with 30 products each. Because there were fewer overall items in some of the categories (such as energy drinks), these items were less frequently drawn in the sample. To fill up the remaining 6 product categories (energy drinks; plant-based milks; edible ices; fresh and frozen meat, poultry, fish and similar; savoury plant-based foods/meat analogues; butter, other fats and oils) we applied the following procedure: We first entered example products from each of the categories into the Open Food Facts search bar and identified relevant tags used by Open Food Facts to identify those products. Using RipGrep, we then filtered for all items which used the relevant tags for each category. Some categories contained multiple tags; all relevant tags were used. Then we used JQ to create a separate CSV file for each of the 6 categories. We imported these CSV files into R and then used the sample function to randomly draw batches of 20 or 30 products from the new datasets. Products were sampled until we had identified 30 products per category.

***Sample size***

We included 30 products per product category, or 660 products in total. We decided on this sample size based on deliberations in the author team, in which we considered the following aspects: feasibility in the time frame of the study and with the available resources; likelihood of including a reasonably broad range and diversity of products in each product category; and the sample size used in comparable projects (in other countries, initial pilot tests of the WHO NPM were conducted on samples of 10-20 products per category, before it was applied to larger samples [5]). We decided that a sample size of 30 products per product category would still be feasible, and more likely to cover the range and diversity of products on the German market than a sample size of 10-20 products per category.

***Data extraction***

Open Food Facts provides product information in two formats: i) as photographs of the packages of products, including the list of ingredients and the nutrient declaration; and ii) as text entered by Open Food Facts contributors (registration as a contributor is open to anyone). We exclusively used the information provided through the photographs of packages. One author (NH, AL, CK, or EO) located each product sampled through the procedure described above in the Open Food Facts database, and then filled our data extraction sheet (see below) with the information provided through the photographs of the ingredient lists and nutrient declarations.

The data extraction sheet included the following items (nutrient data were entered as per 100 g for solid foods, and per 100 ml for liquid foods):

- WHO NPM Product Category
- Product Name (including brand name)

- Open Food Facts URL
- Energy (kcal)
- Total fat (g)
- Saturated fat (g)
- Total sugars (g)
- Salt (g)
- Sodium (g)
- Added sugars (yes/no)
- Non-sugar sweeteners (yes/no)

#### 1.2. Quality control procedures

***Assignment of products to their respective WHO NPM product category***

While assigning products to products categories, authors working on this task (NH, AL, CK, and EO) highlighted all products for which the categorization did not seem fully clear. A second author (CK and PvP) double-checked the assignment of all products to their WHO NPM product category and highlighted products warranting further discussion due to possible ambiguities around their categorization. All highlighted products were then discussed within the team of authors working on the product categorization (NH, AL, CK, EO, and PvP) until a decision was reached. Where necessary, the GFSA codes provided by the WHO NPM manual were checked to provide further guidance. Some products that were at first incorrectly categorized were moved to the appropriate category. This includes two *Kräuterquark* (a traditional German dairy product), which we had originally assigned to category 7 (yogurt, sour milk, cream and similar foods), and which we subsequently reassiged to category 8 (cheese). This also includes a number of further products which had been misclassified and which we either reassigned to the new, correct product category, or which we excluded if the new category already included 30 products (these products are shown in the list of excluded products in section 7). We also took note of any challenges encountered during product assignment (see section 1.3 for a detailed description of these).

***Data extraction***

For 6% of all products (the first and fifteenth item in each category), a second reviewer (CK) checked that the nutrient and ingredient information had been extracted correctly from the Open Food Facts database. No errors were detected.

***Comparison of data provided by Open Food Facts with data provided through the websites of manufacturers and online retailers***

Open Food Facts is based on the collaboration of volunteers who upload product information to the database. Therefore, it is possible that some of the data entered is no longer up to date. For each product, Open Food Facts provides information on the data source: the name of the person that added the product and which persons collaborated on that page, date of last edit, and in some cases the date of the last check. However, it cannot be traced what was changed during the last edit. Hence, one author (CK) checked for 10% of all sampled products if the data provided by the Open Food Facts database matched with nutrient and ingredients data provided on the website of manufacturers or (if no manufacturer website could be identified) on the website of online retailers. For 15 products (23%), we were not able to identify any current information (mostly products from discounters that only provide information on a small part of their product portfolio online). Regarding the presence of added sugars and non-sugar sweeteners, no differences were found between the two data sources. Regarding the nutrient content data, a deviation of 10% or more was found for 42 of 462 data points (9.1%). This includes 25 data points (5.4%) for which the website of the manufacturer or retailer stated a higher value than the one shown on the product photograph provided by Open Food Facts, and 17 data points (3.7%) for which the value provided by the manufacturer or retailer was lower than the one extracted from Open Food Facts.

#### 1.3. Assignment of products to product categories

We used the data provided by the WHO NPM manual to assign products to product categories [5], as described in the Methods section of the main manuscript and in section 1.1 above. Challenges encountered during this process are described in Table s1 below.

**Table s1: Challenges in assigning products to the WHO NPM product categories**

| Issue | Description |
|---|---|
| **Cookie bars** | Category 1 of the WHO NPM includes chocolate, sugar confectionery, energy bars and granola and cereal-type bars, while category 2 includes cakes, sweet biscuits, pastries and other sweet bakery wares (including cakes and cookies covered by chocolate). Based on the descriptions provided by the WHO NPM, we found it difficult to decide whether cookie bars that contain a biscuit component covered with chocolate should be assigned to category 1 or 2. For our analysis, we included them in category 1. |
| **Water-based products that are not used for drinking, but for cooking/baking** | Category 4d) of the WHO NPM includes soft drinks, bottled waters and other drinks (including water-based flavoured drinks), while category 18 includes sauces, dips and dressings. We found it difficult to decide whether products that are technically fruit juices mixed with water, but which are more likely to be used as dressing than as beverage (such as lemon juice mixed with water, marketed as a dressing and cooking ingredient) should be placed in category 4d) or 18. In our analysis, we included them in category 18. |
| **Plant-based yogurt substitutes** | Category 7 of the WHO NPM includes yogurts, including yogurt products containing additional ingredients, as well as yogurt substitutes, while category 2 includes tofu- and plant-based desserts. We found it difficult to decide whether fermented soy products marketed as yogurt substitutes (but which are sometimes called “plant-based desserts” on their labels) should be assigned to category 2 or 7. In our analysis, we considered them to be yogurt substitutes and included them in category 7. |
| **Savoury plant-based spreads** | Category 8 of the WHO NPM includes cheese and cheese analogues, including plant-based, dairy-free cheese and spreads, while category 18 includes sauces, dips and dressings, including guacamole and bean-based dips such as hummus. We found it difficult to decide whether plant-based savoury products that can be used both as spreads and as dips (and that are often marketed as suitable for both purposes) should be placed in category 8 or 18. In our analysis, we included them in category 18. |
| **Potato-based, pasta-like products** | Category 16 includes ready-to-eat potato products, while category 12 includes fresh and dried pasta, including products with the GSFA codes 6.4.21 and 6.4.2 (which include “pasta-like products”). We found it difficult to decide whether pasta-like products made primarily out of potatoes (such |

| Table s1: Challenges in assigning products to the WHO NPM product categories | |
|---|---|
| **Issue** | **Description** |
| | as Gnocchi, or *Schupfnudeln*, a German speciality comparable to Gnocchi) should be placed in category 12 or 6. In our analysis, we included Gnocchi in category 6 (as they are typically marketed as pasta-like product), and Schnupfnudeln in category 16 (as they are a ready-to-eat potato product). |
| **Plain seeds** | WHO NPM category 3 includes savoury snacks, including plain nuts and seeds, while category 6 includes breakfast cereals, including plain oats for preparing muesli made with oats and unsalted nuts and seeds. We found it difficult to decide whether plain seeds that are not typically used as snacks, but rather as ingredient for muesli or similar dishes (such as plain flax, chia, or sesame seeds) should be included in category 3 or 6. In our analysis, we included them in category 3, as they are clearly plain seeds. |
| **Foods for special dietary uses** | The WHO NPM explicitly excludes foods for special dietary uses, defined as “foods that are specially processed or formulated to satisfy particular dietary requirements due to a particular physical or physiological condition and/or specific disease or disorder and which are presented as such”. We found it difficult to decide whether products with a reduced fat, sugar, and/or energy content labelled and advertised as suitable for weight-control or weight-loss purposes (but which are otherwise similar to products not marketed as such) should be excluded or not. In our analysis, we excluded products covered by EU Regulation No 609/2023 (Regulation on Food for Specific Groups), including food for special medical purposes and total diet and meal replacement products for weight control, but not other products carrying nutrition and health claims related to weight loss or maintenance (regulated by EU Regulation No 1924/2006). |
| ***Kräuterquark*** | WHO NPM category 7 (yogurt and cream) includes fromage frais, which is similar to the German Quark. However, category 8 (cheese) also includes unripened cheese e.g., cream cheese, mozzarella, ricotta and cottage cheese, which are also products also similar to Quark. Therefore, we found it difficult to decide whether *Kräuterquark* should be put in category 7 or 8. For our analysis, we considered these products to be cheese and included them in category 8. |

#### 2. Detailed description of the product categories defined by the WHO NPM

**Table s2: Detailed description of the product categories defined by the WHO NPM (Source: [5])**

| No. | WHO product category | Examples | Energy (kcal) | Total fat (g) | Saturated fat (g) | Total sugars (g) | Sodium (g) | Added sugars (g) | Non-sugar sweeteners (g) |
|---|---|---|---|---|---|---|---|---|---|
| **1** | Chocolate and sugar confectionery, energy bars, sweet toppings and desserts | Chocolate confectionery<br>Sugar confectionery (including jellies and boiled sweets; chewing-gum and bubble gum; caramels; liquorice sweets, marzipan sweets)<br>Granola and cereal-type bars<br>Spreadable chocolate and other sweet sandwich toppings<br>Nut butters. (e.g., peanut butter)<br>Honey | | | | | | 0 | 0 |
| **2** | Cakes, sweet biscuits and pastries; other sweet bakery wares; and dry mixes for making such | Cookies/sweet biscuits<br>Cakes and sponges<br>Pies and pastries<br>Baked and cooked desserts<br>Pancakes, waffles and French toast<br>Scones and soda bread<br>Dry mixes for making such<br>Tofu- and other plant-based desserts | | 3 | | | 0.1 | 0 | 0 |
| **3** | Savoury snacks | Crackers/savoury biscuits<br>Nuts, seeds and kernels (including popcorn, nuts, peanuts and seeds (plain or seasoned with salt or flavoured)<br>Potato, vegetable and grain chips<br>Extruded snacks<br>Savoury pretzels | | | | | 0.1 | 0 | 0 |
| **4.1** | Juices | 100% fruit and vegetable juices (including juices reconstituted from concentrate)<br>Smoothies (including smoothies containing yogurt but in which yogurt is not the main ingredient) | | | | 0 | | | 0 |
| **4.2** | Dairy milk drinks | Dairy milks (both sweetened and unsweetened)<br>Milkshakes and coffees containing dairy milk (in which the main constituent is dairy milk) | | 3 | | | | 0 | 0 |
| **4.3** | Plant-based milks | Plant-based milks (both sweetened and unsweetened).<br>Milkshakes and coffees containing plant-based milks (in which the main constituent is plant-based milk) | | 3 | | | | 0 | 0 |

| Table s2: Detailed description of the product categories defined by the WHO NPM (Source: [5]) | | | | | | | | | |
|---|---|---|---|---|---|---|---|---|---|
| **No.** | **WHO product category** | **Examples** | **Energy (kcal)** | **Total fat (g)** | **Saturated fat (g)** | **Total sugars (g)** | **Sodium (g)** | **Added sugars (g)** | **Non-sugar sweeteners (g)** |
| **4.4** | Energy drinks | Beverages containing caffeine or other stimulants such as guarana, taurine, lucuronolactone and vitamins | | | | | | 0 | 0 |
| **4.5** | Soft drinks, bottled waters and other drinks | Water-based flavoured drinks (carbonated and still)<br>Fruit and vegetable nectars<br>Waters (including mineral waters)<br>Coffee, coffee substitutes, tea, herbal infusions and other hot cereal and grain beverages | | | | | | 0 | 0 |
| **5** | Edible ices | Dairy and plant-based ice creams<br>Water-based ices (including sorbets)<br>Frozen yogurts | | 3 | | | 0.1 | 0 | 0 |
| **6** | Breakfast cereals | Minimally processed breakfast cereals (such as steel-cut, rolled or instant oats for preparing oatmeal and muesli; includes porridge mix and hot instant cereals)<br>Highly processed breakfast cereals (including shredded, flaked, puffed and extruded cereals, including granola.) | | 17 | | 12.5 | 0.5 | | |
| **7** | Yogurt, sour milk, cream and similar foods | Yogurt and sour milks (including kefir; buttermilk; flavoured sour, fermented milk and drinking yogurt; fromage frais; cheese-based and other yogurt substitutes)<br>Yogurt products containing additional ingredients (including fruit and muesli)<br>Cream | | 3 | 1 | 12.5 | 0.1 | | |
| **8** | Cheese | Hard, medium and soft cheeses (unripened and ripened).<br>Processed cheeses (including cheese spreads) | | 17 | | | 0.5 | | |
| **9** | Ready-made and convenience foods and composite dishes | Tinned composite foods (including meat balls in sauce and baked beans)<br>Pasta, noodles and rice or grains with sauce or seasoned<br>Pizza and pizza snacks<br>Sandwiches and wraps (including hamburgers and hot dogs)<br>Prepared salads<br>Ready-to-eat meals composed of a combination of carbohydrate and either vegetable or meat, or all three combined<br>Soups (ready-to eat, tinned and refrigerated and dry and concentrated) | | 17 | 6 | 12.5 | 0.5 | | |

| Table s2: Detailed description of the product categories defined by the WHO NPM (Source: [5]) | | | | | | | | | |
|---|---|---|---|---|---|---|---|---|---|
| **No.** | **WHO product category** | **Examples** | **Energy (kcal)** | **Total fat (g)** | **Saturated fat (g)** | **Total sugars (g)** | **Sodium (g)** | **Added sugars (g)** | **Non-sugar sweeteners (g)** |
| **10** | Butter, other fats and oils | Butter, butter blends, margarine and oil-based spreads<br>Vegetable oils | | | 21 | | 0.5 | | |
| **11** | Bread, bread products and crisp breads | Sweet and raisin breads (including brioche)<br>Leavened bread (including breads made with all types of cereal flours, e.g., white or whole-grain wheat, spelt and rye)<br>Flatbreads | | 17 | | 12.5 | 0.5 | | |
| **12** | Fresh or dried pasta, rice and grains | Fresh or dried pasta and noodles<br>Rice and grains | | 17 | | 12.5 | 0.5 | | |
| **13** | Fresh and frozen meat, poultry, fish and similar | Fresh and frozen meat, poultry, game and fish<br>Eggs | | 17 | | | | | |
| **14** | Processed meat, poultry, fish and similar | Processed fish and seafood products (including tinned, raw and non-heat-treated; e.g.,, tinned tuna, smoked fish and fish fingers)<br>Processed meat, poultry, game and preparations (including tinned, raw, heat- and non-heat-treated, e.g., ham, burgers, sausages and breaded meat products) | | 17 | | | 0.5 | | |
| **15** | Fresh and frozen fruit, vegetables and legumes | Fresh and frozen fruit, vegetables without additional ingredients (including starch vegetables, roots and tubers)<br>Fresh and frozen legumes without additional ingredients. | Permitted | | | | | | |
| **16** | Processed fruit and vegetables | Tinned, pickled, dried, battered and breaded vegetables and legumes<br>Tinned, dried and pickled fruits<br>Fruit and vegetable pouches | | 3 | | 12.5 | 0.5 | 0 | |
| **17** | Savoury plant-based foods/ meat analogues | Tofu and tempeh<br>Meat analogues (including “veggie” burgers) | | 17 | | | 0.5 | 0 | 0 |
| **18** | Sauces, dips and dressings | Stock cubes<br>Cooking sauces (including pasta sauces)<br>Dips and dipping sauces<br>Salad dressings<br>Condiments (including tomato ketchups) | | 17 | | | 0.5 | 0 | 0 |

#### 3. Additional analyses

The following tables show the results of our analyses conducted with the original WHO NPM without the adaptations proposed by Germany's Federal Ministry for Food and Agriculture, which we report in the main manuscript.

**Table s3: Share of products below nutrient and ingredient thresholds: WHO NPM without adaptations**

| | Energy | Total fat | Saturated fat | Total sugar | Sodium | Added sugars | Non-sugar sweeteners |
|---|---|---|---|---|---|---|---|
| **Median across product categories (IQR)** | **80% (80%-80%)** | **77% (35%-95%)** | **73% (45%-87%)** | **77% (63%-98%)** | **65% (40%-96%)** | **37% (10%-58%)** | **100% (97%-100%)** |
| **1 Confectionery** | | | | | | 3% | 97% |
| **2 Cakes and cookies** | | 7% | | | 40% | 0% | 100% |
| **3 Savoury snacks** | | | | | 53% | 77% | 97% |
| **4.1 Juices** | | | | 0% | | | 100% |
| **4.2 Dairy milk drinks** | | 37% | | | | 80% | 100% |
| **4.3 Plant-based milks** | | 93% | | | | 73% | 100% |
| **4.4 Energy drinks** | | | | | | 30% | 53% |
| **4.5 Soft drinks, bottled water and other drinks** | | | | | | 37% | 80% |
| **5 Ice cream** | | 17% | | | 100% | 0% | 97% |
| **6 Breakfast cereals** | | 100% | | 60% | 97% | | |
| **7 Yogurt and cream** | | 33% | 17% | 77% | 97% | | |
| **8 Cheese** | | 27% | | | 33% | | |
| **9 Ready-made and convenience foods** | 80% | 97% | 100% | 100% | 70% | | |
| **10 Butter, other fats and oils** | | | 73% | | 100% | | |
| **11 Bread** | | 100% | | 97% | 60% | | |
| **12 Pasta and grains** | | 100% | | 100% | 93% | | |
| **13 Fresh and frozen meat, fish and eggs** | | 93% | | | | | |
| **14 Processed meat and fish** | | 53% | | | 17% | | |
| **15 Fresh and frozen fruit and vegetables** | | | | | | | |
| **16 Processed fruit and vegetables** | | 77% | | 67% | 77% | 43% | |
| **17 Savoury plant-based foods** | | 93% | | | 20% | 37% | 100% |
| **18 Sauces, dips and dressings** | | 67% | | | 40% | 17% | 100% |

Abbreviations: WHO NPM: World Health Organization Regional Office for Europe Nutrient Profiling Model [5].

| Table s4: Median content relative to threshold: WHO NPM without adaptations* | | | | | | | |
|---|---|---|---|---|---|---|---|
| | **Energy (%)** | **Total fat (%)** | **Saturated fat (%)** | **Total sugar (%)** | **Sodium (%)** | **Added sugars**** | **Non-sugar sweeteners**** |
| **Median across product categories (IQR)** | **68% (68%-68%)** | **65% (36%-118%)** | **82% (46%-84%)** | **29% (20%-44%)** | **66% (39%-127%)** | **n.a.** | **n.a.** |
| **1 Confectionery** | | | | | | n.a. | n.a. |
| **2 Cakes and cookies** | | 700% | | | 157% | n.a. | n.a. |
| **3 Savoury snacks** | | | | | 38% | n.a. | n.a. |
| **4.1 Juices** | | | | | | n.a. | n.a. |
| **4.2 Dairy milk drinks** | | 118% | | | | n.a. | n.a. |
| **4.3 Plant-based milks** | | 47% | | | | n.a. | n.a. |
| **4.4 Energy drinks** | | | | | | n.a. | n.a. |
| **4.5 Soft drinks, bottled water and other drinks** | | | | | | n.a. | n.a. |
| **5 Ice cream** | | 367% | | | 48% | n.a. | n.a. |
| **6 Breakfast cereals** | | 34% | | 84% | 4% | | |
| **7 Yogurt and cream** | | 117% | 82% | 36% | 40% | | |
| **8 Cheese** | | 156% | | | 132% | | |
| **9 Ready-made and convenience foods** | 68% | 39% | 11% | 19% | 83% | | |
| **10 Butter, other fats and oils** | | | 86% | | 0% | | |
| **11 Bread** | | 20% | | 21% | 96% | | |
| **12 Pasta and grains** | | 10% | | 16% | 1% | | |
| **13 Fresh and frozen meat, fish and eggs** | | 65% | | | | | |
| **14 Processed meat and fish** | | 91% | | | 184% | | |
| **15 Fresh and frozen fruit and vegetables** | | | | | | | |
| **16 Processed fruit and vegetables** | | 17% | | 46% | 47% | n.a. | |
| **17 Savoury plant-based foods** | | 74% | | | 132% | n.a. | n.a. |
| **18 Sauces, dips and dressings** | | 50% | | | 112% | n.a. | n.a. |

Abbreviations: WHO NPM: World Health Organization Regional Office for Europe Nutrient Profiling Model[5]. *For added sugars and non-sugar sweeteners, the threshold defined by the adapted WHO NPM is 0; it is therefore not possible to state the content of these nutrients relative to the threshold.

| Table s5: Share of products exceeding a defined number of thresholds: WHO NPM without adaptations | | | | | |
|---|---|---|---|---|---|
| | Mean number of thresholds exceeded | Share of products exceeding a defined number of thresholds | | | |
| | | 0 | 1 | 2 | 3 |
| **Median across product categories (IQR)** | **1.2 (1.0-1.7)** | **20% (3%-59%)** | **28% (18%-43%)** | **7% (0%-46%)** | **0% (0%-3%)** |
| **1 Confectionery** | 1.0 | 0% | 100% | 0% | 0% |
| **2 Cakes and cookies** | 2.5 | 0% | 7% | 33% | 60% |
| **3 Savoury snacks** | 1.6 | 53% | 23% | 20% | 3% |
| **4.1 Juices** | 1.0 | 0% | 100% | 0% | 0% |
| **4.2 Dairy milk drinks** | 1.0 | 20% | 77% | 3% | 0% |
| **4.3 Plant-based milks** | 1.1 | 70% | 27% | 3% | 0% |
| **4.4 Energy drinks** | 1.2 | 0% | 83% | 17% | 0% |
| **4.5 Soft drinks, bottled water and other drinks** | 1.1 | 23% | 70% | 7% | 0% |
| **5 Ice cream** | 1.9 | 0% | 17% | 80% | 3% |
| **6 Breakfast cereals** | 1.0 | 57% | 43% | 0% | 0% |
| **7 Yogurt and cream** | 2.0 | 13% | 10% | 63% | 13% |
| **8 Cheese** | 1.8 | 20% | 20% | 60% | 0% |
| **9 Ready-made and convenience foods** | 1.3 | 60% | 30% | 7% | 3% |
| **10 Butter, other fats and oils** | 1.0 | 73% | 27% | 0% | 0% |
| **11 Bread** | 1.0 | 57% | 43% | 0% | 0% |
| **12 Pasta and grains** | 1.0 | 93% | 7% | 0% | 0% |
| **13 Fresh and frozen meat, fish and eggs** | 1.0 | 93% | 7% | 0% | 0% |
| **14 Processed meat and fish** | 1.5 | 13% | 43% | 43% | 0% |
| **15 Fresh and frozen fruit and vegetables** | n.a. | 100% | 0% | 0% | 0% |
| **16 Processed fruit and vegetables** | 1.7 | 20% | 27% | 50% | 3% |
| **17 Savoury plant-based foods** | 1.7 | 10% | 37% | 47% | 7% |
| **18 Sauces, dips and dressings** | 1.8 | 0% | 37% | 50% | 13% |
| Abbreviations: WHO NPM: World Health Organization Regional Office for Europe Nutrient Profiling Model [5]. | | | | | |

#### 4. Differences between protocol and manuscript

This study is based on a protocol developed and prospectively registered and published online through the Open Science Framework (registration https://doi.org/10.17605/OSF.IO/BJEVC) before data were analysed [6]. In the following, we describe differences between the protocol and the manuscript:

- In the sampling process (described in section 1), we found that six of the 22 product categories filled up slowly, as the full sample of products from the Open Food Facts database contained relatively few products included in these six categories. For these six product categories, we therefore modified the sampling procedure described in our protocol by filtering for products that were tagged in Open Food Facts with terms related to the respective product categories;

we then drew a random sample from this sub-sample of tagged products, and assigned these successively to the remaining product categories until these were all filled with 30 products.

- In our protocol, we had planned two quality control measures (namely that a second author would double-check for all products that they were assigned to the correct product category, and for 6% of all products that the data had been extracted correctly from the Open Food Facts database). To ensure quality, we added the two additional quality control measures described in the methods section of the manuscript, and in section 1 of the supplementary material (i.e. the discussion in the team of authors of all products for which the assignment to the correct product category was found challenging, and the validation with information from manufacturer and retailer websites).
- In our protocol, we anticipated that we would calculate the mean threshold exceedance of products (i.e. the mean value by which products that exceed a specific nutrient threshold exceed the respective threshold). While conducting our analysis, we found that in many product categories, the number of products exceeding each indidivual threshold was low, resulting in a very small n for many of these analyses. We therefore decided to calculate the mean nutrient content of products instead.

### 5. Code

#### 5.1. Code for the random sampling (for R)

```
library(rjson)
library(dplyr)

datafood <- read.csv("openfoodfacts-germany.csv")

sample_n(datafood, 100)
sample_n(datafood, 50)
sample_n(datafood, 50)

secondatafood <- read.csv("openfoodfacts-front-photo.csv")
sample_n(secondatafood, 30)

secondatafood %>% select(product_name, brand) %>% sample_n(30)
sample_n(secondatafood, 30)

datafood2 <- read.csv("openfoodfacts-front-photo.csv")
sample_n(datafood2, 50)

datafoodenergy <- read.csv("openfoodfacts-energy-drinks.csv", header=
FALSE)
sample_n(datafoodenergy, 20)

datafoodfette <- read.csv("openfoodfacts-fette.csv", header =FALSE)
sample_n(datafoodfette, 30)

datafoodmilkreplace <- read.csv("openfoodfacts-milk-replacements.csv" ,
header = FALSE)
sample_n(datafoodmilkreplace, 30)

datafoodspeiseeis <- read.csv("openfoodfacts-speiseeis.csv" , header =
FALSE)
sample_n(datafoodspeiseeis, 30)
```

```
datafoodfakemeat <- read.csv("openfoodfacts-meat-replacements.csv" , header
= FALSE)
sample_n(datafoodfakemeat, 30)

datafoodmeat <- read.csv("openfoodfacts-fleisch.csv", header = FALSE)

sample_n(datafoodmeat, 100)
sample_n(datafoodmeat, 20)
sample_n(datafoodmeat, 50)
sample_n(secondatafood, 50)
```

#### 5.2. Code for the statistical analyses (for R)

```
library(dplyr)
library(tidyverse)
library(openxlsx)

npm_data <- read.csv('… /npm_data.csv')

##categorize data correctly
npm_data <- mutate(npm_data,
                   who_cat=as.factor(who_cat),
                   added_sugars=as.factor(added_sugars),
                   sweeteners=as.factor(sweeteners)
)

# rename who_cat to food_cat and establish numbers as who_cat
npm_data <- mutate (npm_data, food_cat=who_cat,
      who_cat=ifelse(
      who_cat=="Confectionery",1,ifelse(
      who_cat=="Cakes / cookies",2,ifelse(
      who_cat=="Snacks",3,ifelse(
      who_cat=="Juice",4.1,ifelse(
      who_cat=="Dairy milk",4.2,ifelse(
      who_cat=="Plant milk",4.3,ifelse(
      who_cat=="Energy drinks",4.4,ifelse(
      who_cat=="Soft drinks / water",4.5,ifelse(
      who_cat=="Ice cream",5,ifelse(
      who_cat=="Cereals",6,ifelse(
      who_cat=="Yogurt / cream",7,ifelse(
      who_cat=="Cheese",8,ifelse(
      who_cat=="Convenience food",9,ifelse(
      who_cat=="Fats",10,ifelse(
      who_cat=="Bread",11,ifelse(
      who_cat=="Pasta / grains",12,ifelse(
      who_cat=="Meat / fish / eggs",13,ifelse(
      who_cat=="Processed meat / fish",14,ifelse(
      who_cat=="Fruit / vegetables",15,ifelse(
      who_cat=="Processed fruit / vegetables",16,ifelse(
      who_cat=="Plant-based food",17,ifelse(
      who_cat=="Sauces",18,0))))))))))))))))))))))
      )

#establish reformulation variables: These can be used for reformulation
scenarios
en_reform <- c(1)
fat_reform <- c(1)
sat_fat_reform <- c(1)
total_sugars_reform <- c(1)
sodium_reform <- c(1)
```

```
#In this version, the lines for juice (4.1) and milk (4.2) thresholds are
commented out. They can be uncommented for the WHO NPM without adaptations

######################################################
##share of products within energy threshold
######################################################

npm_data <- mutate(npm_data,
     en_thresh=ifelse(who_cat==9,1,0),
     exc_en_thresh=ifelse (who_cat== 9 & en*en_reform > 225,1,0))

en <- npm_data%>%
  filter(en_thresh==1)%>%
  summarise(percent_within_en = sum(exc_en_thresh==0)/sum(en_thresh==1))

#by food category
en_cat <- npm_data%>%
  filter(en_thresh==1)%>%
  group_by(who_cat,food_cat)%>%
  summarise(percent_within_en = sum(exc_en_thresh==0)/sum(en_thresh==1))

#############################################################
##Share of products within the total FAT threshold
#############################################################

npm_data <- mutate(npm_data,
   fat_thresh= ifelse(
     who_cat==2|
       #who_cat==4.2|
       who_cat==4.3|
       who_cat==5|
       who_cat==6|
       who_cat==7|
       who_cat==8|
       who_cat==9|
       who_cat==11|
       who_cat==12|
       who_cat==13|
       who_cat==14|
       who_cat==16|
       who_cat==17|
       who_cat==18,1,0),
   exc_fat_thresh=ifelse (
     (who_cat==2 & total_fat*fat_reform > 3)|
       #(who_cat==4.2 & total_fat*fat_reform > 3)|
       (who_cat==4.3 & total_fat*fat_reform > 3)|
       (who_cat==5 & total_fat*fat_reform > 3)|
       (who_cat==6 & total_fat*fat_reform > 17)|
       (who_cat==7 & total_fat*fat_reform > 3)|
       (who_cat==8 & total_fat*fat_reform > 17)|
       (who_cat==9 & total_fat*fat_reform > 17)|
       (who_cat==11 & total_fat*fat_reform > 17)|
       (who_cat==12 & total_fat*fat_reform > 17)|
       (who_cat==13 & total_fat*fat_reform > 17)|
       (who_cat==14 & total_fat*fat_reform > 17)|
       (who_cat==16 & total_fat*fat_reform > 3)|
       (who_cat==17 & total_fat*fat_reform > 17)|
       (who_cat==18 & total_fat*fat_reform > 17),1,0)
)
```

```
fat <- npm_data%>%
  filter(fat_thresh==1)%>%
  summarise(percent_within_fat = sum(exc_fat_thresh==0)/sum(fat_thresh==1))

#by food category
fat_cat <- npm_data %>%
  filter(fat_thresh==1)%>%
  group_by(who_cat,food_cat)%>%
  summarise(percent_within_fat = sum(exc_fat_thresh==0)/sum(fat_thresh==1))

######################################################################
##Share of products within the total SATURATED FAT threshold
######################################################################

npm_data <- mutate(npm_data,
   sat_fat_thresh= ifelse(
     who_cat== 7 |
       who_cat== 9 |
       who_cat== 10,1,0),
   exc_sat_fat_thresh = ifelse (
     (who_cat== 7 & sat_fat*sat_fat_reform > 1)|
       (who_cat== 9 & sat_fat*sat_fat_reform > 6)|
       (who_cat== 10 & sat_fat*sat_fat_reform > 21),1,0)
)

sat_fat <- npm_data %>%
  filter(sat_fat_thresh==1)%>%
  summarise(percent_within_sat_fat =
sum(exc_sat_fat_thresh==0)/sum(sat_fat_thresh==1))

#by food category
sat_fat_cat <- npm_data %>%
  filter(sat_fat_thresh==1)%>%
  group_by(who_cat,food_cat)%>%
  summarise(percent_within_sat_fat =
sum(exc_sat_fat_thresh==0)/sum(sat_fat_thresh==1))

###############################################################
##Share of products within the total SUGAR threshold
###############################################################

npm_data <- mutate(npm_data,
    sugar_thresh= ifelse(
      #who_cat==4.1|
      who_cat==6|
       who_cat==7|
       who_cat==9|
       who_cat==11|
       who_cat==12|
       who_cat==16,1,0),
      exc_sugar_thresh=ifelse (
      #(who_cat==4.1 & total_sugars*total_sugars_reform > 0)|
      (who_cat==6 & total_sugars*total_sugars_reform > 12.5)|
       (who_cat==7 & total_sugars*total_sugars_reform > 12.5)|
       (who_cat==9 & total_sugars*total_sugars_reform > 12.5)|
       (who_cat==11 & total_sugars*total_sugars_reform > 12.5)|
       (who_cat==12 & total_sugars*total_sugars_reform > 12.5)|
       (who_cat==16 & total_sugars*total_sugars_reform > 12.5),1,0))

sugar <- npm_data %>%
```

```
  filter(sugar_thresh==1)%>%
  summarise(percent_within_sugar =
sum(exc_sugar_thresh==0)/sum(sugar_thresh==1))

#by food category
sugar_cat <- npm_data %>%
  filter(sugar_thresh==1)%>%
  group_by(who_cat,food_cat)%>%
  summarise(percent_within_sugar =
sum(exc_sugar_thresh==0)/sum(sugar_thresh==1))

################################################################
##Share of products within the total SODIUM threshold
################################################################

npm_data <- mutate(npm_data,
   sodium_thresh=ifelse(
     who_cat==2|
       who_cat==3|
       who_cat==5|
       who_cat==6|
       who_cat==7|
       who_cat==8|
       who_cat==9|
       who_cat==10|
       who_cat==11|
       who_cat==12|
       who_cat==14|
       who_cat==16|
       who_cat==17|
       who_cat==18,1,0),
   exc_sodium_thresh = ifelse (
     (who_cat==2 & sodium*sodium_reform > 0.1)|
       (who_cat==3 & sodium*sodium_reform > 0.1)|
       (who_cat==5 & sodium*sodium_reform > 0.1)|
       (who_cat==6 & sodium*sodium_reform > 0.5)|
       (who_cat==7 & sodium*sodium_reform > 0.1)|
       (who_cat==8 & sodium*sodium_reform > 0.5)|
       (who_cat==9 & sodium*sodium_reform > 0.5)|
       (who_cat==10 & sodium*sodium_reform > 0.5)|
       (who_cat==11 & sodium*sodium_reform > 0.5)|
       (who_cat==12 & sodium*sodium_reform > 0.5)|
       (who_cat==14 & sodium*sodium_reform > 0.5)|
       (who_cat==16 & sodium*sodium_reform > 0.5)|
       (who_cat==17 & sodium*sodium_reform > 0.5)|
       (who_cat==18 & sodium*sodium_reform > 0.5),1,0)
)

sodium <- npm_data%>%
  filter(sodium_thresh==1)%>%
  summarise(percent_within_sodium =
sum(exc_sodium_thresh==0)/sum(sodium_thresh==1))

#by food category
sodium_cat <- npm_data%>%
  filter(sodium_thresh==1)%>%
  group_by(who_cat,food_cat)%>%
  summarise(percent_within_sodium =
sum(exc_sodium_thresh==0)/sum(sodium_thresh==1))
```

```
##############################################################
##Share of products within the ADDED SUGAR threshold
##############################################################

npm_data <- mutate(npm_data,
   added_sugars_thresh= ifelse(
     who_cat==1|
       who_cat==2|
       who_cat==3|
       who_cat==4.2|
       who_cat==4.3|
       who_cat==4.4|
       who_cat==4.5|
       who_cat==5|
       who_cat==16|
       who_cat==17|
       who_cat==18,1,0),
   exc_added_sugars_thresh = ifelse (
     (who_cat==1 & added_sugars=="yes")|
       (who_cat==2 & added_sugars=="yes")|
       (who_cat==3 & added_sugars=="yes")|
       (who_cat==4.2 & added_sugars=="yes")|
       (who_cat==4.3 & added_sugars=="yes")|
       (who_cat==4.4 & added_sugars=="yes")|
       (who_cat==4.5 & added_sugars=="yes")|
       (who_cat==5 & added_sugars=="yes")|
       (who_cat==16 & added_sugars=="yes")|
       (who_cat==17 & added_sugars=="yes")|
       (who_cat==18 & added_sugars=="yes"),1,0)
)

added_sugars <- npm_data%>%
  filter(added_sugars_thresh==1)%>%
  summarise(percent_within_added_sugars =
sum(exc_added_sugars_thresh==0)/sum(added_sugars_thresh==1))

#by food category
added_sugars_cat <- npm_data%>%
  filter(added_sugars_thresh==1)%>%
  group_by(who_cat,food_cat)%>%
  summarise(percent_within_added_sugars =
sum(exc_added_sugars_thresh==0)/sum(added_sugars_thresh==1))

#############################################################
##Share of products within the sweeteners threshold
#############################################################

npm_data <- mutate(npm_data,
   sweeteners_thresh= ifelse(
     who_cat==1|
       who_cat==2|
       who_cat==3|
       who_cat==4.1|
       who_cat==4.2|
       who_cat==4.3|
       who_cat==4.4|
       who_cat==4.5|
       who_cat==5|
       who_cat==17|
       who_cat==18,1,0),
   exc_sweeteners_thresh = ifelse(
```

```
      (who_cat==1 & sweeteners=="yes")|
        (who_cat==2 & sweeteners=="yes")|
        (who_cat==3 & sweeteners=="yes")|
        (who_cat==4.1 & sweeteners=="yes")|
        (who_cat==4.2 & sweeteners=="yes")|
        (who_cat==4.3 & sweeteners=="yes")|
        (who_cat==4.4 & sweeteners=="yes")|
        (who_cat==4.5 & sweeteners=="yes")|
        (who_cat==5 & sweeteners=="yes")|
        (who_cat==17 & sweeteners=="yes")|
        (who_cat==18 & sweeteners=="yes"),1,0)
)

sweeteners <- npm_data%>%
  filter(sweeteners_thresh==1)%>%
  summarise(percent_within_sweeteners =
sum(exc_sweeteners_thresh==0)/sum(sweeteners_thresh==1))

#by food category
sweeteners_cat <- npm_data%>%
  filter(sweeteners_thresh==1)%>%
  group_by(who_cat,food_cat)%>%
  summarise(percent_within_sweeteners =
sum(exc_sweeteners_thresh==0)/sum(sweeteners_thresh==1))

####################################################################
#### CREATE RESULTS TABLE###########################################
####################################################################

# share of items that are within threshold by food category
list_shares=list(
  select(en_cat,'who_cat','percent_within_en'),
  select(fat_cat,'who_cat','percent_within_fat'),
  select(sat_fat_cat,'who_cat','percent_within_sat_fat'),
  select(sugar_cat,'who_cat','percent_within_sugar'),
  select(sodium_cat,'who_cat','percent_within_sodium'),
  select(added_sugars_cat,'who_cat','percent_within_added_sugars'),
  select(sweeteners_cat,'who_cat','percent_within_sweeteners'))

shares_cat <- list_shares %>% reduce(full_join,by='who_cat') %>%
arrange(who_cat)

#Median and IQR of shares of different food categories that are within each
threshold
median_shares_within <- shares_cat%>%
  group_by()%>%
  summarise(
    en = median(percent_within_en,na.rm=TRUE),
    fat = median(percent_within_fat,na.rm=TRUE),
    sat_fat = median(percent_within_sat_fat,na.rm=TRUE),
    sugar = median(percent_within_sugar,na.rm=TRUE),
    sodium = median(percent_within_sodium,na.rm=TRUE),
    added_sugars = median(percent_within_added_sugars,na.rm=TRUE),
    sweeteners = median(percent_within_sweeteners,na.rm=TRUE))

iqr1_shares_within_threshold <- shares_cat%>%
  group_by()%>%
  summarise(en=quantile(percent_within_en,1/4,na.rm=TRUE),
    fat=quantile(percent_within_fat,1/4,na.rm=TRUE),
    sat_fat=quantile(percent_within_sat_fat,1/4,na.rm=TRUE),
```

```
    sugar=quantile(percent_within_sugar,1/4,na.rm=TRUE),
    sodium=quantile(percent_within_sodium,1/4,na.rm=TRUE),
    added_sugars=quantile(percent_within_added_sugars,1/4,na.rm=TRUE),
    sweeteners=quantile(percent_within_sweeteners,1/4,na.rm=TRUE))

iqr3_shares_within_threshold <- shares_cat%>%
  group_by()%>%
  summarise(en=quantile(percent_within_en,3/4,na.rm=TRUE),
    fat=quantile(percent_within_fat,3/4,na.rm=TRUE),
    sat_fat=quantile(percent_within_sat_fat,3/4,na.rm=TRUE),
    sugar=quantile(percent_within_sugar,3/4,na.rm=TRUE),
    sodium=quantile(percent_within_sodium,3/4,na.rm=TRUE),
    added_sugars=quantile(percent_within_added_sugars,3/4,na.rm=TRUE),
    sweeteners=quantile(percent_within_sweeteners,3/4,na.rm=TRUE))

#create table with median and IQR
iqr_threshold <- rbind(median_shares_within,
                       iqr1_shares_within_threshold,
                       iqr3_shares_within_threshold)
row.names(iqr_threshold)<- c("median","iqr1","iqr3")

#########################################################################
## count the number of items that exceeded 1, 2, or 3 thresholds
#########################################################################

#add the number of thresholds exceeded for each item:
npm_data <- mutate(npm_data,
                   number_thresh_exc= exc_en_thresh + exc_fat_thresh +
exc_sat_fat_thresh + exc_sugar_thresh + exc_sodium_thresh +
exc_added_sugars_thresh + exc_sweeteners_thresh)

#by food category: count how many items exceeed 0,1,2,3 thresholds
threshold_cat <- npm_data%>%
  group_by(who_cat,food_cat)%>%
  count(number_thresh_exc)

#take percentage:
threshold_cat <- threshold_cat%>%
  mutate(percentage=n/sum(n))

#reorganize data in table:
threshold_cat <- threshold_cat%>%

pivot_wider(id_cols=food_cat,names_from=number_thresh_exc,values_from=perce
ntage)
colnames(threshold_cat)<-c('food_cat','One','Two','Three','Zero')

threshold_cat[is.na(threshold_cat)] <- 0

#calculate the median and IQR for share of items that exceed 0,1,2, or 3
thresholds
threshold_median_0123 <- threshold_cat%>%
  group_by()%>%
  summarise(
    md_zero=median(Zero,na.rm=TRUE),
    iqr1_zero=quantile(Zero,1/4,na.rm=TRUE),
    iqr3_zero=quantile(Zero,3/4,na.rm=TRUE),
    md_one=median(One,na.rm=TRUE),
    iqr1_one=quantile(One,1/4,na.rm=TRUE),
    iqr3_one=quantile(One,3/4,na.rm=TRUE),
    md_two=median(Two,na.rm=TRUE),
```

```
    iqr1_two=quantile(Two,1/4,na.rm=TRUE),
    iqr3_two=quantile(Two,3/4,na.rm=TRUE),
    md_three=median(Three,na.rm=TRUE),
    iqr1_three=quantile(Three,1/4,na.rm=TRUE),
    iqr3_three=quantile(Three,3/4,na.rm=TRUE))

####################################################
## Calculate median and mean of thresholds exceeded
####################################################

threshold_median <- npm_data%>%
  filter(number_thresh_exc>0)%>%
  summarise(med = median(number_thresh_exc),mn=mean(number_thresh_exc))

threshold_median_cat <- npm_data%>%
  filter(number_thresh_exc>0)%>%
  group_by(who_cat,food_cat)%>%
  summarise(med = median(number_thresh_exc),
    mn=mean(number_thresh_exc),
    quart1=quantile(number_thresh_exc,1/4),
    quart3=quantile(number_thresh_exc,3/4))

#add the median of these to threshold_median_0123
threshold_median_0123 <-
  mutate(threshold_median_0123,
     md_total=median(threshold_median_cat$mn),
     iqr1_total=quantile(threshold_median_cat$mn,1/4),
     iqr3_total=quantile(threshold_median_cat$mn,3/4)
  )

#########################################################
## create healthy variable and calculate share of healthy
#########################################################

npm_data <- mutate(npm_data,
   healthy_r= ifelse(number_thresh_exc>0,"unhealthy","healthy"))

# share of healthy by cat
healthy_cat <- npm_data%>%
  group_by(who_cat,food_cat)%>%
  summarise(percent_healthy = sum(healthy_r=="healthy")/n())

# calculate median, ICR of share of healthy
healthy_median <- healthy_cat %>%
  group_by()%>%
  summarise(med = median(percent_healthy),
    mn=mean(percent_healthy),
    quant1=quantile(percent_healthy,1/4),
    quant3=quantile(percent_healthy,3/4))

#transpose healthy_cat for export:
healthy_cat <- as.data.frame(t(healthy_cat))

#######################################
##Energy content relative to threshold
#######################################

npm_data <- mutate(npm_data,

en_content_percent=ifelse(who_cat==9,(en*en_reform)/(225),0))
```

```
#by food category:
en_content_cat <- npm_data %>%
  filter(en_thresh==1)%>%
  group_by(who_cat)%>%
  summarise(rel_median_en = median(en_content_percent))

#median en content:
en_content <- en_content_cat %>%
  summarise(
    md=median(rel_median_en),
    rel_iqr1_en=quantile(rel_median_en,1/4),
    rel_iqr3_en=quantile(rel_median_en,3/4))

####################################
##Fat content relative to threshold
####################################

npm_data <- mutate(npm_data,
  fat_content_percent=ifelse(
    who_cat==2,
    (total_fat*fat_reform)/(3),ifelse(
    #who_cat== 4.2,
    #(total_fat*fat_reform)/(3),ifelse(
    who_cat==4.3,(total_fat*fat_reform)/(3),ifelse(
    who_cat==5,(total_fat*fat_reform)/(3),ifelse(
    who_cat==6,(total_fat*fat_reform)/(17),ifelse(
    who_cat==7,(total_fat*fat_reform)/(3),ifelse(
    who_cat==8,(total_fat*fat_reform)/(17),ifelse(
    who_cat==9,(total_fat*fat_reform)/(17),ifelse(
    who_cat==11,(total_fat*fat_reform)/(17),ifelse(
    who_cat==12,(total_fat*fat_reform)/(17),ifelse(
    who_cat==13,(total_fat*fat_reform)/(17),ifelse(
    who_cat==14,(total_fat*fat_reform)/(17),ifelse(
    who_cat==16,(total_fat*fat_reform)/(3),ifelse(
    who_cat==17,(total_fat*fat_reform)/(17),ifelse(
    who_cat==18,(total_fat*fat_reform)/(17),0))))))))))))))#)

#by food category:
fat_content_cat <- npm_data%>%
  filter(fat_thresh==1)%>%
  group_by(who_cat)%>%
  summarise(rel_median_fat = median(fat_content_percent))

#median fat content:
fat_content <- fat_content_cat%>%
  group_by()%>%
  summarize(
    md=median(rel_median_fat),
    rel_iqr1_fat=quantile(rel_median_fat,1/4),
    rel_iqr3_fat=quantile(rel_median_fat,3/4))

###########################################################################
###
##Sat fat content relative to threshold
###########################################################################
###

npm_data <- mutate(npm_data,
```

```
  sat_fat_content_percent= ifelse(
  who_cat==7,sat_fat*sat_fat_reform/(3),ifelse(
  who_cat==9,sat_fat*sat_fat_reform/(17),ifelse(
  who_cat==10,sat_fat*sat_fat_reform/(17),0))))

#by food category:
sat_fat_content_cat <- npm_data%>%
  filter(sat_fat_thresh==1)%>%
  group_by(who_cat)%>%
  summarise(rel_median_sat_fat=median(sat_fat_content_percent))

#median sat fat content:
sat_fat_content <- sat_fat_content_cat%>%
  summarise(
    md=median(rel_median_sat_fat),
    rel_iqr1_sat_fat=quantile(rel_median_sat_fat,1/4),
    rel_iqr3_sat_fat=quantile(rel_median_sat_fat,3/4))

######################################
##Sugar content relative to threshold
######################################

npm_data <- mutate(npm_data,sugar_content_percent = ifelse (
    #who_cat== 4.1 ,total_sugars*total_sugars_reform/(0),ifelse(
    who_cat== 6,(total_sugars*total_sugars_reform)/(12.5),ifelse(
    who_cat== 7,(total_sugars*total_sugars_reform)/(12.5),ifelse(
    who_cat== 9,(total_sugars*total_sugars_reform)/(12.5),ifelse(
    who_cat== 11,(total_sugars*total_sugars_reform)/(12.5),ifelse(
    who_cat== 12,(total_sugars*total_sugars_reform)/(12.5),ifelse(
    who_cat== 16,(total_sugars*total_sugars_reform)/(12.5),0))))))
)

#by food category:
sugar_content_cat <- npm_data%>%
  filter(sugar_thresh==1)%>%
  group_by(who_cat)%>%
  summarise(rel_median_sugar=median(sugar_content_percent))

#median sugar content:
sugar_content <- sugar_content_cat%>%
  summarise(
    md=median(rel_median_sugar),
    rel_iqr1_sugar=quantile(rel_median_sugar,1/4),
    rel_iqr3_sugar=quantile(rel_median_sugar,3/4))

#######################################
##sodium content relative to threshold
#######################################

npm_data <- mutate(npm_data,
    sodium_content_percent = ifelse (
      who_cat==2,(sodium*sodium_reform)/(0.1),ifelse(
      who_cat==3,(sodium*sodium_reform)/(0.1),ifelse(
      who_cat==5,(sodium*sodium_reform)/(0.1),ifelse(
      who_cat==6,(sodium*sodium_reform)/(0.5),ifelse(
      who_cat==7,(sodium*sodium_reform)/(0.1),ifelse(
      who_cat==8,(sodium*sodium_reform)/(0.5),ifelse(
      who_cat==9,(sodium*sodium_reform)/(0.5),ifelse(
      who_cat==10,(sodium*sodium_reform)/(0.5),ifelse(
      who_cat==11,(sodium*sodium_reform)/(0.5),ifelse(
```

```
      who_cat==12,(sodium*sodium_reform)/(0.5),ifelse(
      who_cat==14,(sodium*sodium_reform)/(0.5),ifelse(
      who_cat==16,(sodium*sodium_reform)/(0.5),ifelse(
      who_cat==17,(sodium*sodium_reform)/(0.5),ifelse(
      who_cat==18,(sodium*sodium_reform)/(0.5),0)))))))))))))
  )

#by food category:
sodium_content_cat <- npm_data%>%
  filter(sodium_thresh==1)%>%
  group_by(who_cat)%>%
  summarise(rel_median_sodium=median(sodium_content_percent))

#median sodium content:
sodium_content <- sodium_content_cat%>%
  summarise(
    md=median(rel_median_sodium),
    rel_iqr1_sodium=quantile(rel_median_sodium,1/4),
    rel_iqr3_sodium=quantile(rel_median_sodium,3/4))

########################################################
#### CREATE RESULTS TABLE for median nutrient content###
########################################################

#median, iqr across all items
content <- matrix(c(
  en_content[1],en_content[2],en_content[3],
  fat_content[1],fat_content[2],fat_content[3],
  sat_fat_content[1],sat_fat_content[2],sat_fat_content[3],
  sugar_content[1],sugar_content[2],sugar_content[3],
  sodium_content[1],sodium_content[2],sodium_content[3]),
  ncol=5, byrow=FALSE)
colnames(content) <- c('en','fat','saturated_fat','sugar','sodium')
rownames(content) <- c('median','iqr1','iqr3')

#by food category:
list_content = list(en_content_cat,
    fat_content_cat,
    sat_fat_content_cat,
    sugar_content_cat,
    sodium_content_cat)

content_cat <- list_content%>%
  reduce(full_join,by='who_cat')%>%
  arrange(who_cat)

###################################
#Create Excel file################
###################################

npm_analysis <- createWorkbook()

# Add sheets to the file

#table 2
addWorksheet(npm_analysis,"healthy_median")
addWorksheet(npm_analysis, "healthy_cat")

#table 3
addWorksheet(npm_analysis,"iqr_threshold")
```

```
addWorksheet(npm_analysis,"shares_cat")

#table 4
addWorksheet(npm_analysis, "content")
addWorksheet(npm_analysis,"content_cat")

#table 5
addWorksheet(npm_analysis,"threshold_median_0123")
addWorksheet(npm_analysis,"threshold_cat")
addWorksheet(npm_analysis,"threshold_median_cat")

# Write data to the sheets

writeData(npm_analysis,1,healthy_median)
writeData(npm_analysis,2,healthy_cat,rowNames=TRUE,colNames=FALSE)

writeData(npm_analysis,3,iqr_threshold,rowNames=TRUE,colNames=TRUE)
writeData(npm_analysis,4,shares_cat,rowNames=FALSE,colNames=TRUE)

writeData(npm_analysis,5,content,rowNames=TRUE,colNames=TRUE)
writeData(npm_analysis,6,content_cat,rowNames=FALSE,colNames=TRUE)

writeData(npm_analysis,7,threshold_median_0123,rowNames=FALSE,colNames=TRUE
)
writeData(npm_analysis,8,threshold_cat,rowNames=FALSE,colNames=TRUE)
writeData(npm_analysis,9,threshold_median_cat,rowNames=FALSE,colNames=TRUE)

# Save the file
saveWorkbook(npm_analysis,"npm_results_bmel.xlsx",overwrite=TRUE)
```

#### 6. Sampling and data extraction guidance sheet

The following guidance sheet was used to ensure consistency among the authors applying the sampling and data extraction procedures described in section 1.

##### Sampling and data extraction procedure

1) Copy and paste the product name (and brand, if listed) from the provided R list (Sheet 6) and search for it in the search bar of https://de.openfoodfacts.org/ Make sure the language for the page is in Deutsch. (Top left corner of the website). See Exclusion Criteria below to check if the product meets criteria.

2) When searching for products for which you have the name AND brand, select the product which exactly matches the product and brand name in the search results. If there are multiple entries that match the exact search term (name and brand), select the first product in the list. If no item in the first page of results exactly matches the name and brand in your search term, follow instructions in 2c.

    a) If the search term only yields one result, check the product for exclusion criteria and then continue the procedure in Step 3.

    b) If the product that exactly matches the name and brand does not contain all the necessary info AND if there are multiple search results for that product name and brand, then return to the first page of search results and follow instructions in 2c.

c) If there is no brand given: select the first product that appears in the search result (it is ok if the name does not exactly match what you had, *as long as it is essentially the same product*). If the data for that item is incomplete (if it meets exclusion criteria), return to the search results and select the second item in the list. Continue this pattern until you reach an item that has all necessary data. *If none of the search results for that product contain all the necessary info, then exclude the product and enter it in Sheet 5.*

d) *If the ingredient list is only in a language other than German:* return to the search results and select the second item in the list that most closely matches your search term. If there are no other items to select, exclude the product.

e) Please note that fresh fruits and vegetables will not contain an ingredient list or nutrition facts panel. These are still to be included as long as they have a picture.

f) Fresh meats (like chicken breast or ground beef) will usually also not contain a nutrition facts panel. These are to be included.

g) Whole grains (like oats, bulgur, lentils) *with no added ingredients* will not contain an ingredient list. These are to be included.

h) According to policy, products with only one ingredient do not need to list an ingredient list. These in general should be included if other criteria are met.

3) Once you have selected the product, decide which WHO food/beverage category it belongs to. Copy and paste the page title–product name and brand (please exclude the size measurement)--into the Google Sheet (Sheet 3) under the correct category.
4) Copy and paste the URL of the product page into the Google Sheet.
5) Fill out the remaining columns (D-J) for the product in the Google Sheet by reading the images of the ingredient list (to check for added sugars and non-sugar sweeteners) and the nutrition facts panel. Please do not use the embedded table of nutrition facts listed on the website. Read the image of the nutrition facts panel.
6) For added sugars and non-sugar sweeteners: please just note whether these ingredients are listed. It is a binary choice in Google Sheets: yes or no. For all products, please write yes/no under both added sugars and non-sugar sweeteners.
7) Total sugars are listed under “davon Zucker.”
8) Columns K and L are automated. So please do not enter data into these columns.
9) If you cannot confidently sort an item into one of the 22 food/beverage categories, enter the product name and brand at the end of Sheet 3.

##### Exclusion Criteria

Once you search for the item in the Open Food Facts database, check to make sure the product does not meet any exclusion criteria, as listed below:

1) Exclude products with no product image
2) Exclude products with no image of the nutrition facts panel
3) Exclude products with no image of the ingredient list
4) Exclude products where the ingredient list AND nutrition facts panel are presented only in a language other than German.

5) Exclude products where the ingredient list is only presented in a language other than German.
6) Exclude products which are outside the scope of the model (listed on page 15 of the manual)
7) Exclude non-food or non-beverage items (ex. Cleaning supplies or medicines)
8) Exclude alcoholic beverages
9) Exclude plain spices, flour, vinegar, protein powders

For products that meet the exclusion criteria, please enter the item in Sheet 5 of the Google Sheet and select a reason for exclusion from the dropdown menu.

**Common Names of Added Sugars (From WHO Manual)**: "[A]dded sugars have many different names, including brown sugar, cane juice, corn syrup, dextrose, fructose, fruit nectars, glucose, high-fructose corn syrup, honey, lactose, malt syrup, maltose, maple syrup, molasses, raw sugar and sucrose."

#### 7. STROBE-nut reporting guideline

The STROBE-nut reporting guideline checklist is available online at: https://www.strobe-nut.ugent.be/content/recommendations

For a detailed description of the STROBE and STROBE-nut reporting guideline see von Elm 2007 and Lachat 2016 [7, 8].

Page numbers with a preceding s (s1, s2, etc.) refer to the supplementary material.

**Table s6: STROBE-nut Checklist**

| Nr | Item | STROBE recommendations | STROBE-nut | Page Nr |
|---|---|---|---|---|
| 1 | **Title and abstract** | (a) Indicate the study's design with a commonly used term in the title or the abstract.<br>(b) Provide in the abstract an informative and balanced summary of what was done and what was found. | **nut-1** State the dietary/nutritional assessment method(s) used in the title, abstract, or keywords. | 1-2 |
| | **Introduction** | | | |
| 2 | Background rationale | Explain the scientific background and rationale for the investigation being reported. | | 3-4 |
| 3 | Objectives | State specific objectives, including any pre-specified hypotheses. | | 4 |
| | **Methods** | | | |

| Table s6: STROBE-nut Checklist | | | | |
|---|---|---|---|---|
| **Nr** | **Item** | **STROBE recommendations** | **STROBE-nut** | **Page Nr** |
| 4 | Study design | Present key elements of study design early in the paper. | | 4 |
| 5 | Settings | Describe the setting, locations, and relevant dates, including periods of recruitment, exposure, follow-up, and data collection. | **nut-5** Describe any characteristics of the study settings that might affect the dietary intake or nutritional status of the participants, if applicable. | 4-5<br>s1-s5 |
| 6 | Participants | a) Cohort study—Give the eligibility criteria, and the sources and methods of selection of participants. Describe methods of follow-up.<br>Case-control study—Give the eligibility criteria, and the sources and methods of case ascertainment and control selection. Give the rationale for the choice of cases and controls.<br>Cross-sectional study—Give the eligibility criteria, and the sources and methods of selection of participants.<br>(b) Cohort study—For matched studies, give matching criteria and number of exposed and unexposed.<br>Case-control study—For matched studies, give matching criteria and the number of controls per case. | **nut-6** Report particular dietary, physiological or nutritional characteristics that were considered when selecting the target population. | n.a. |
| 7 | Variables | Clearly define all outcomes, exposures, predictors, potential confounders, and effect modifiers. Give diagnostic criteria, if applicable. | **nut-7.1** Clearly define foods, food groups, nutrients, or other food components.<br>**nut-7.2** When using dietary patterns or indices, describe the methods to obtain them and their nutritional properties. | 4-5<br>s1-s5 |

| Table s6: STROBE-nut Checklist | | | | |
|---|---|---|---|---|
| **Nr** | **Item** | **STROBE recommendations** | **STROBE-nut** | **Page Nr** |
| 8 | Data sources - measurements | For each variable of interest, give sources of data and details of methods of assessment (measurement). Describe comparability of assessment methods if there is more than one group. | **nut-8.1** Describe the dietary assessment method(s), e.g., portion size estimation, number of days and items recorded, how it was developed and administered, and how quality was assured. Report if and how supplement intake was assessed.<br>**nut-8.2** Describe and justify food composition data used. Explain the procedure to match food composition with consumption data. Describe the use of conversion factors, if applicable.<br>**nut-8.3** Describe the nutrient requirements, recommendations, or dietary guidelines and the evaluation approach used to compare intake with the dietary reference values, if applicable.<br>**nut-8.4** When using nutritional biomarkers, additionally use the STROBE Extension for Molecular Epidemiology (STROBE-ME). Report the type of biomarkers used and their usefulness as dietary exposure markers.<br>**nut-8.5** Describe the assessment of nondietary data (e.g., nutritional status and influencing factors) and timing of the assessment of these variables in relation to dietary assessment.<br>**nut-8.6** Report on the validity of the dietary or nutritional assessment methods and any internal or external validation used in the study, if applicable. | 4-5<br>s1-s5<br>s24-s26 |
| 9 | Bias | Describe any efforts to address potential sources of bias. | **nut-9** Report how bias in dietary or nutritional assessment was addressed, e.g., misreporting, | 4-5<br>s1-s5 |

| Table s6: STROBE-nut Checklist | | | | |
|---|---|---|---|---|
| **Nr** | **Item** | **STROBE recommendations** | **STROBE-nut** | **Page Nr** |
| | | | changes in habits as a result of being measured, or data imputation from other sources | s24-s26 |
| 10 | Study Size | Explain how the study size was arrived at. | | s2 |
| 11 | Quantitative variables | Explain how quantitative variables were handled in the analyses. If applicable, describe which groupings were chosen and why. | **nut-11** Explain categorization of dietary/nutritional data (e.g., use of N-tiles and handling of nonconsumers) and the choice of reference category, if applicable. | 4-5<br>S1-s5 |
| 12 | Statistical Methods | (a) Describe all statistical methods, including those used to control for confounding<br>(b) Describe any methods used to examine subgroups and interactions.<br>(c) Explain how missing data were addressed.<br>(d) Cohort study—If applicable, explain how loss to follow-up was addressed.<br>Case-control study—If applicable, explain how matching of cases and controls was addressed.<br>Cross-sectional study—If applicable, describe analytical methods taking account of sampling strategy.<br>(e) Describe any sensitivity analyses. | **nut-12.1** Describe any statistical method used to combine dietary or nutritional data, if applicable.<br>**nut-12.2** Describe and justify the method for energy adjustments, intake modeling, and use of weighting factors, if applicable.<br>**nut-12.3** Report any adjustments for measurement error, i.e,. from a validity or calibration study. | 4.5<br>s1-s5 |
| | **Results** | | | |

| Table s6: STROBE-nut Checklist | | | | |
|---|---|---|---|---|
| **Nr** | **Item** | **STROBE recommendations** | **STROBE-nut** | **Page Nr** |
| 13 | Participants | (a) Report the numbers of individuals at each stage of the study—e.g., numbers potentially eligible, examined for eligibility, confirmed eligible, included in the study, completing follow-up, and analyzed.<br>(b) Give reasons for non-participation at each stage.<br>(c) Consider use of a flow diagram. | **nut-13** Report the number of individuals excluded based on missing, incomplete or implausible dietary/nutritional data. | n.a. |
| 14 | Descriptive data | (a) Give characteristics of study participants (e.g., demographic, clinical, social) and information on exposures and potential confounders<br>(b) Indicate the number of participants with missing data for each variable of interest<br>(c) Cohort study—Summarize follow-up time (e.g., average and total amount) | **nut-14** Give the distribution of participant characteristics across the exposure variables if applicable. Specify if food consumption of total population or consumers only were used to obtain results. | n.a. |
| 15 | Outcome data | Cohort study—Report numbers of outcome events or summary measures over time.<br>Case-control study—Report numbers in each exposure category, or summary measures of exposure.<br>Cross-sectional study—Report numbers of outcome events or summary measures. | | 7-11 |
| 16 | Main results | (a) Give unadjusted estimates and, if applicable, confounder-adjusted estimates and their precision (e.g., 95% confidence interval).<br>Make clear which confounders were adjusted for and why they were included. | **nut-16** Specify if nutrient intakes are reported with or without inclusion of dietary supplement intake, if applicable. | n.a. |

| Table s6: STROBE-nut Checklist | | | | |
|---|---|---|---|---|
| **Nr** | **Item** | **STROBE recommendations** | **STROBE-nut** | **Page Nr** |
| | | (b) Report category boundaries when continuous variables were categorized.<br>(c) If relevant, consider translating estimates of relative risk into absolute risk for a meaningful time period. | | |
| 17 | Other analyses | Report other analyses done—e.g., analyses of subgroups and interactions and sensitivity analyses. | **nut-17** Report any sensitivity analysis (e.g., exclusion of misreporters or outliers) and data imputation, if applicable. | |
| | **Discussion** | | | |
| 18 | Key results | Summarize key results with reference to study objectives. | | 13 |
| 19 | Limitation | Discuss limitations of the study, taking into account sources of potential bias or imprecision. Discuss both direction and magnitude of any potential bias. | **nut-19** Describe the main limitations of the data sources and assessment methods used and implications for the interpretation of the findings. | 13-14 |
| 20 | Interpretation | Give a cautious overall interpretation of results considering objectives, limitations, multiplicity of analyses, results from similar studies, and other relevant evidence. | **nut-20** Report the nutritional relevance of the findings, given the complexity of diet or nutrition as an exposure. | 14-15 |

| Table s6: STROBE-nut Checklist | | | | |
|---|---|---|---|---|
| **Nr** | **Item** | **STROBE recommendations** | **STROBE-nut** | **Page Nr** |
| 21 | Generalizability | Discuss the generalizability (external validity) of the study results. | | 14 |
| | **Other information** | | | |
| 22 | Funding | Give the source of funding and the role of the funders for the present study and, if applicable, for the original study on which the present article is based. | | 16 |
| | *Ethics* | | **nut-22.1** Describe the procedure for consent and study approval from ethics committee(s). | 15 |
| | *Supplementary material* | | **nut-22.2** Provide data collection tools and data as online material or explain how they can be accessed. | s1-s38 |

#### 8. List of products excluded after reassignment

This table shows products that were reassigned to a different product category after discussions in the team of authors (see section 1.2), and which were excluded from the analysis because the new product category already contained 30 products.

**Table s7: List of products excluded after reassignment**

| WHO Category | Product Name | URL | Energy (kcal) | Total fat (g) | Saturated fat (g) | Total sugars (g) | Salt (g) | Sodium (g) | Added sugars (g) | Non-sugar sweeteners (g) |
|---|---|---|---|---|---|---|---|---|---|---|
| **2. Cakes / cookies** | Butter Spritzringe - knackig Et zart | https://de.openfoodfacts.org/produkt/4335896084848/butter-spritzringe-knackig-et-zart | 532 | 30 | 21 | 23 | 0,33 | 0,132 | yes | no |
| | Knusprige Waffelbecher - Biscotto - 60 g | https://de.openfoodfacts.org/produkt/29012145/knusprige-waffelbecher-biscotto | 496 | 24 | 22 | 20 | 0,4 | 0,16 | yes | no |
| **3. Snacks** | Bio Goldleinsamen, geschrotet - Alnatura | https://de.openfoodfacts.org/produkt/4104420010826/bio-goldleinsamen-geschrotet-alnatura | 471 | 30,9 | 3 | 0 | 0,15 | 0,06 | no | no |
| | Flohsamenschalen - Coop | https://de.openfoodfacts.org/produkt/7624841573573/flohsamenschalen-coop | 184 | 0 | 0 | 0 | 0,2 | 0,08 | no | no |
| **4.2 Dairy milk** | Lait entier - Migros | https://de.openfoodfacts.org/produkt/7613404603079/lait-entier-migros | 64 | 3,6 | 2,2 | 5 | 0,1 | 0,04 | no | no |
| **4.3. Plant milk** | Bio Regeneration Drink - Green Force | https://de.openfoodfacts.org/produkt/4260498850013/bio-regeneration-drink-green-force | 43 | 0,7 | 0,3 | 4,1 | 0,2 | 0,08 | yes | no |
| | Schoko mandel - koawach | https://de.openfoodfacts.org/produkt/4260407951923/schoko-mandel-koawach | 94 | 1,9 | 0,5 | 12 | 0,09 | 0,036 | yes | no |
| **4d. Soft drinks / water** | Heißer Apfel-Ingwer - Rabenhorst | https://de.openfoodfacts.org/produkt/4004191001248/heißer-apfel-ingwer-rabenhorst | 66 | 0,5 | 0,1 | 16 | 0,03 | 0,012 | no | no |

| Table s7: List of products excluded after reassignment | | | | | | | | | | |
|---|---|---|---|---|---|---|---|---|---|---|
| **WHO Category** | **Product Name** | **URL** | **Energy (kcal)** | **Total fat (g)** | **Saturated fat (g)** | **Total sugars (g)** | **Salt (g)** | **Sodium (g)** | **Added sugars (g)** | **Non-sugar sweeteners (g)** |
| | Holunderblüte Blütensirup - Yo - | https://de.openfoodfacts.org/produkt/9001400004200/blütensirup-holunderblüte-yo | 38 | 0,5 | 0,1 | 9,3 | 0,01 | 0,004 | yes | no |
| **5. Ice cream** | Yoghurt & Raspberry - Lagnese | https://de.openfoodfacts.org/produkt/8711327392014/yoghurt-raspberry-lagnese | 318 | 20 | 14 | 29 | 0,07 | 0,028 | yes | no |
| **6. Cereals** | Dinkelherzen - Enerbio | https://de.openfoodfacts.org/produkt/4305615678672/dinkelherzen-enerbio | 494 | 24 | 14 | 32 | 0,07 | 0,028 | yes | no |
| | Porridge WellMix Balance | https://de.openfoodfacts.org/produkt/4305615601458/porridge-wellmix-balance | 117 | 2,7 | 0,7 | 2,2 | 0,19 | 0,076 | yes | no |
| **7. Yogurt / cream** | Pflanzlich aus Kokosmilch - Kirsche - Mövenpick | https://de.openfoodfacts.org/produkt/7720727766777/pflanzlich-aus-kokosmilch-kirsche-mövenpick | 127 | 8 | 7,5 | 8,3 | 0,07 | 0,028 | yes | no |
| **8. Cheese** | Bergkäse mittel | https://de.openfoodfacts.org/produkt/2700002002528/bergkäse-mittel | 422 | 34 | 23 | 0 | 1,5 | 0,6 | no | no |
| **9. Convenience food** | Tortellini Ricotta & Spinaci - Rana | https://de.openfoodfacts.org/produkt/8001665127561/tortellini-ricotta-spinaci-rana | 261 | 8,2 | 2,6 | 4,9 | 1 | 0,4 | no | no |
| | Spinat-Ricotta-Tortelloni - Steinhaus | https://de.openfoodfacts.org/produkt/4009337842815/spinat-ricotta-tortelloni-steinhaus | 226 | 7,5 | 3 | 1,5 | 1 | 0,4 | yes | no |
| | Ravioli alla pizzaiola - Pasta Nuova | https://de.openfoodfacts.org/produkt/4011849500503/ravioli-alla-pizzaiola-pasta-nuova | 266 | 4,4 | 1,5 | 2,4 | 0,76 | 0,304 | no | no |
| **11. Bread** | Dürüm Tortilla Waps | https://de.openfoodfacts.org/produkt/8437011503732/dürüm-tortilla-waps | 312 | 7 | 3,7 | 1,8 | 1,3 | 0,52 | no | no |

| Table s7: List of products excluded after reassignment | | | | | | | | | | |
|---|---|---|---|---|---|---|---|---|---|---|
| **WHO Category** | **Product Name** | **URL** | **Energy (kcal)** | **Total fat (g)** | **Saturated fat (g)** | **Total sugars (g)** | **Salt (g)** | **Sodium (g)** | **Added sugars (g)** | **Non-sugar sweeteners (g)** |
| **14. Processed meat / fish** | Ei + Bacon - chef select | https://de.openfoodfacts.org/produkt/4056489580829/ei-bacon-chef-select | 279 | 24,7 | 3 | 1,8 | 2,16 | 0,864 | yes | no |
| | Matjessalat - ALMARE Seafood | https://de.openfoodfacts.org/produkt/4047247018517/matjessalat-almare-seafood | 276 | 26 | 3,4 | 1,6 | 2,5 | 1 | yes | no |
| | Alaska Seelachs - Sea Gold | https://de.openfoodfacts.org/produkt/4316268481762/alaska-seelachs-sea-gold | 101 | 4,9 | 0,6 | 0 | 9,4 | 3,76 | no | no |
| | Salami tradizionale - Edeka | https://de.openfoodfacts.org/produkt/4311501448458/salami-tradizionale-edeka | 344 | 25,4 | 8,5 | 0,1 | 4 | 1,6 | yes | no |
| **15. Fruit / vegetables** | Bio Rispentomaten - Gut Bio | https://de.openfoodfacts.org/produkt/4061458240628/bio-rispentomaten-gut-bio | | | | | | | | |
| **16. Processed fruit / vegetables** | Sonnen Fruchte - Aprikose Zentis | https://de.openfoodfacts.org/produkt/4002575514469/sonnen-fruchte-aprikose-zentis | 226 | 0,1 | 0,1 | 54 | 0,02 | 0,008 | yes | no |
| | Kartoffel-Schupfnudeln (Fach Svana) - Bürger | https://de.openfoodfacts.org/produkt/4075600033891/kartoffel-schupfnudeln-fach-svana-bürger | 183 | 1,6 | 0,3 | 5,1 | 0,96 | 0,384 | no | no |
| **18. Sauces** | Gewürze - BioBio - Zirtronello - Zitrone - BioBio, BioBio (NETTO) | https://de.openfoodfacts.org/produkt/42381402/gewürze-biobio-zirtronello-zitrone | 23 | 0,5 | 0 | 0,3 | 0,03 | 0,012 | no | no |
